# Biallelic Variants in *KMO* Cause a Novel Form of Congenital NAD Deficiency

**DOI:** 10.64898/2026.08.24.26360911

**Authors:** Nathalie M. Aceves-Ewing, Nanbing Li-Villarreal, Xiaohui Li, Seema R. Lalani, Jill A. Rosenfeld, Varduhi Petrosyan, Aleksandar Milosavljevic, Angelina Gaspero, Denise G. Lanza, Audrey E. Christiansen, Amrit Koirala, Abu Hena Mostafa Kamal, Nagireddy Putluri, Cristian Coarfa, Bao Tran, Philip L. Lorenzi, Lin Tan, Charul Gijavanekar, Sarah H. Elsea, Emily Lawrence, Hartmut Cuny, Sally L. Dunwoodie, Pengfei Liu, Haonan Zhouyao, Tara L. Rasmussen, Mary E. Dickinson, Carlos A. Bacino, Brendan Lee, Ronit Marom, Undiagnosed Diseases Network, BCM Center for Precision Medicine Models, Jason D. Heaney, Chih-Wei Hsu, Lindsay C. Burrage

## Abstract

Congenital NAD deficiency disorder (CNDD) is a gene × environment disorder caused by disruptions of the kynurenine pathway. To date, CNDD has been associated with biallelic variants in three kynurenine pathway genes: *KYNU, HAAO,* and *NADSYN1*. We identified two sisters with congenital anomalies overlapping with CNDD who have biallelic variants in a gene encoding a different kynurenine pathway enzyme, *KMO*. The surviving child also has elevated levels of metabolites upstream of KMO with low NAD^+^ levels in plasma, suggesting that KMO deficiency is a novel CNDD. To explore the pathogenicity of KMO deficiency, we generated a global *Kmo* knockout mouse model (*Kmo^-/-^*) and utilized dietary interventions to better model human gene × environment interactions. Although *Kmo^-/-^*mice are viable and fertile on typical breeder chow, they exhibit elevated serum kynurenine and are functionally vitamin B3-dependent. Under conditions of limited maternal vitamin B3 intake, a greater proportion of *Kmo^-/-^* embryos develop congenital anomalies and have significantly lower NAD^+^ levels than *Kmo^+/-^* littermates. Exploratory untargeted metabolomics performed in *Kmo^-/-^* embryos suggested that NAD^+^ deficiency may perturb the pyrimidine, purine, and pentose phosphate pathways. These findings establish KMO deficiency as a new cause of CNDD and highlight a critical gene × environment interaction influencing NAD metabolism and congenital anomalies.

## Introduction

NAD^+^ and NADH, collectively NAD, are molecules that are essential for a wide range of biological processes. NAD serves as a cofactor for the electron transport chain, energy metabolism, fatty acid oxidation, DNA repair, genomic stability, post-translational modifications, and other processes crucial for homeostasis^1–4^. NAD is synthesized via several pathways, including the evolutionarily conserved kynurenine pathway, which converts dietary tryptophan (TRP) to NAD^+^ through various enzymatic reactions and degrades up to 95% of dietary TRP (**Figure 1)**^1,5–7^. In addition to the kynurenine pathway, the Preiss-Handler and salvage pathways utilize dietary sources of vitamin B3 (such as nicotinic acid and nicotinamide) to synthesize NAD^+8,9^.

**Figure 1.**
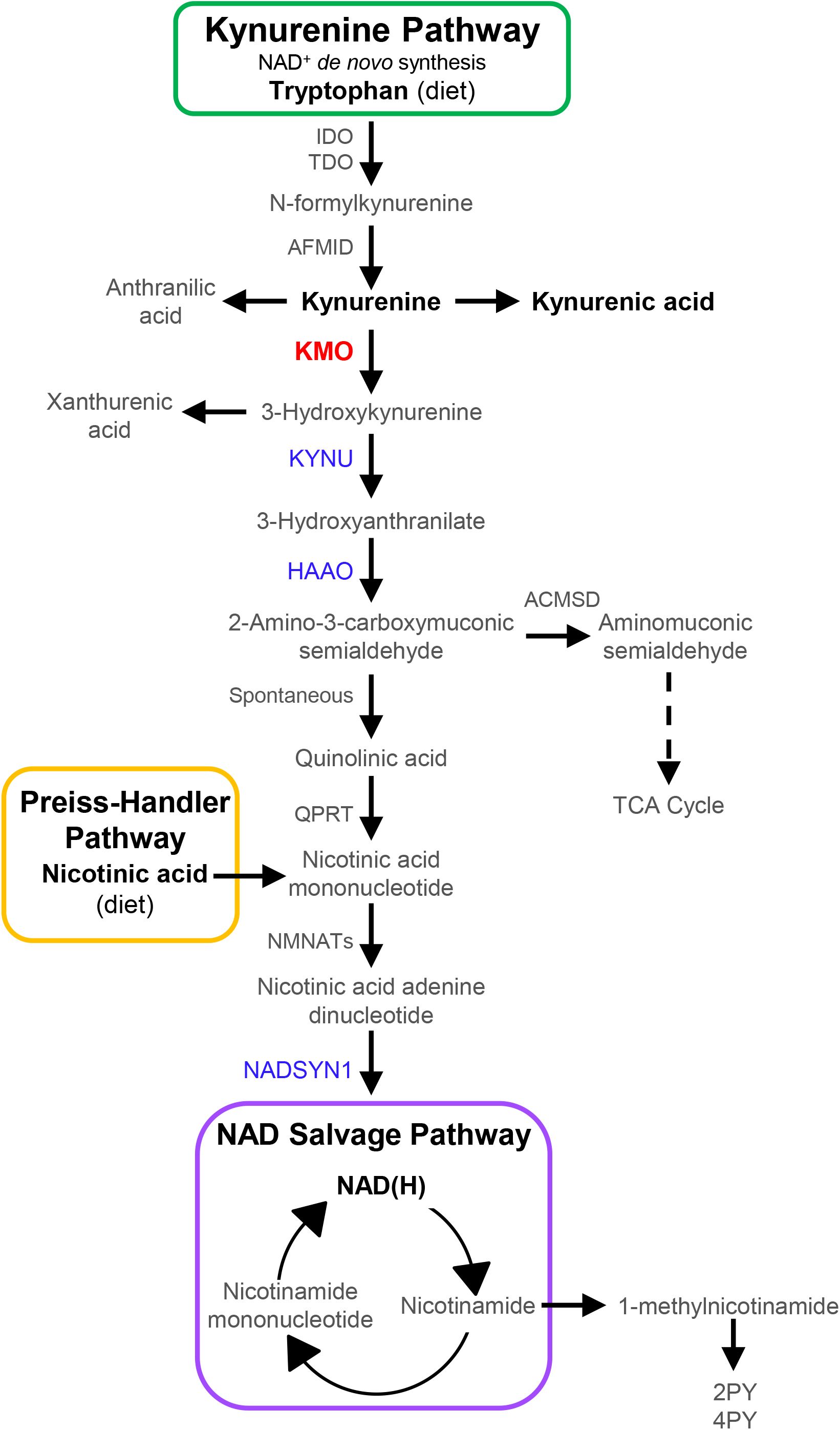
The NAD biosynthesis pathways. Enzymes previously associated with CNDD/VCRL syndrome are in blue.

Congenital deficiency of one of three kynurenine pathway enzymes has been linked to a recognizable human phenotype associated with multiple congenital malformations, recurrent pregnancy loss, and sometimes low NAD, referred to as congenital NAD deficiency disorder (CNDD) or vertebral, cardiac, renal, and limb defects syndrome (VCRL1-3)^10–22^. Thus far, CNDD has been associated with biallelic loss-of-function variants in one of three genes encoding enzymes in the kynurenine pathway: *KYNU, HAAO,* and *NADSYN1* (MIM # 617661, 617660, 618845; **Figure 1**)^10–21^. Individuals with CNDD present with heterogeneous phenotypes that vary in severity, including short stature, short limbs, vertebral, cardiac, renal, and/or limb anomalies, and sometimes neurodevelopmental phenotypes^10–21^. Similarly, mouse embryos with a deletion of *Kynu*, *Haao*, or *Nadsyn1* are at increased risk for embryonic lethality and/or congenital anomalies in the setting of a diet low in NAD precursors^10,12,23,24^. A diet with sufficient NAD precursors, however, prevents congenital anomalies in all embryos, regardless of genotype, indicating that CNDD is a disorder influenced by gene × environment interactions in mice and suggesting that a similar interaction may impact human development^10,12,23,24^.

In this study, we report sisters with biallelic kynurenine 3-monooxygenase (*KMO*) variants and a constellation of congenital anomalies that overlap with CNDD^10,19,25^. *KMO* encodes kynurenine 3-monooxygenase (KMO), an enzyme within the kynurenine pathway that catalyzes the conversion of kynurenine to 3-hydroxykynurenine, the substrate of KYNU (**Figure 1**)^26–28^. We used clinically available metabolomic studies in the family and a global *Kmo* knockout mouse model to demonstrate that KMO deficiency is a novel CNDD that increases the risk for congenital anomalies, some of which may be preventable with vitamin B3 supplementation. Moreover, we used exploratory untargeted metabolomics in developing mouse embryos to identify pathways disrupted during embryogenesis when KMO is deficient.

## Results

### Case Report

Two sisters presented with multiple congenital anomalies, including cardiac abnormalities, vertebral anomalies, and short stature. One sister passed away secondary to the congenital anomalies (II-1 in **Figure 2**). An autopsy confirmed the cardiac and vertebral anomalies as well as short stature and further identified a brain anomaly. Chromosome microarray (Baylor Genetics, CMA-HR+SNP, V10.2) and proband exome sequencing were nondiagnostic. In addition to several cardiac and vertebral anomalies, the proband (II-2 in **Figure 2**) was found to have small kidneys, unilateral hearing loss associated with an abnormal ear, and learning disabilities. A chromosomal microarray (Baylor Genetics, CMA-HR+SNP, V11.2) and trio exome sequencing (analyzed with the deceased sister’s exome data) for the proband were nondiagnostic. *As per medRxiv policy, the detailed case histories for the participants and their images have been removed. To obtain more information, please contact the authors*.

**Figure 2.**
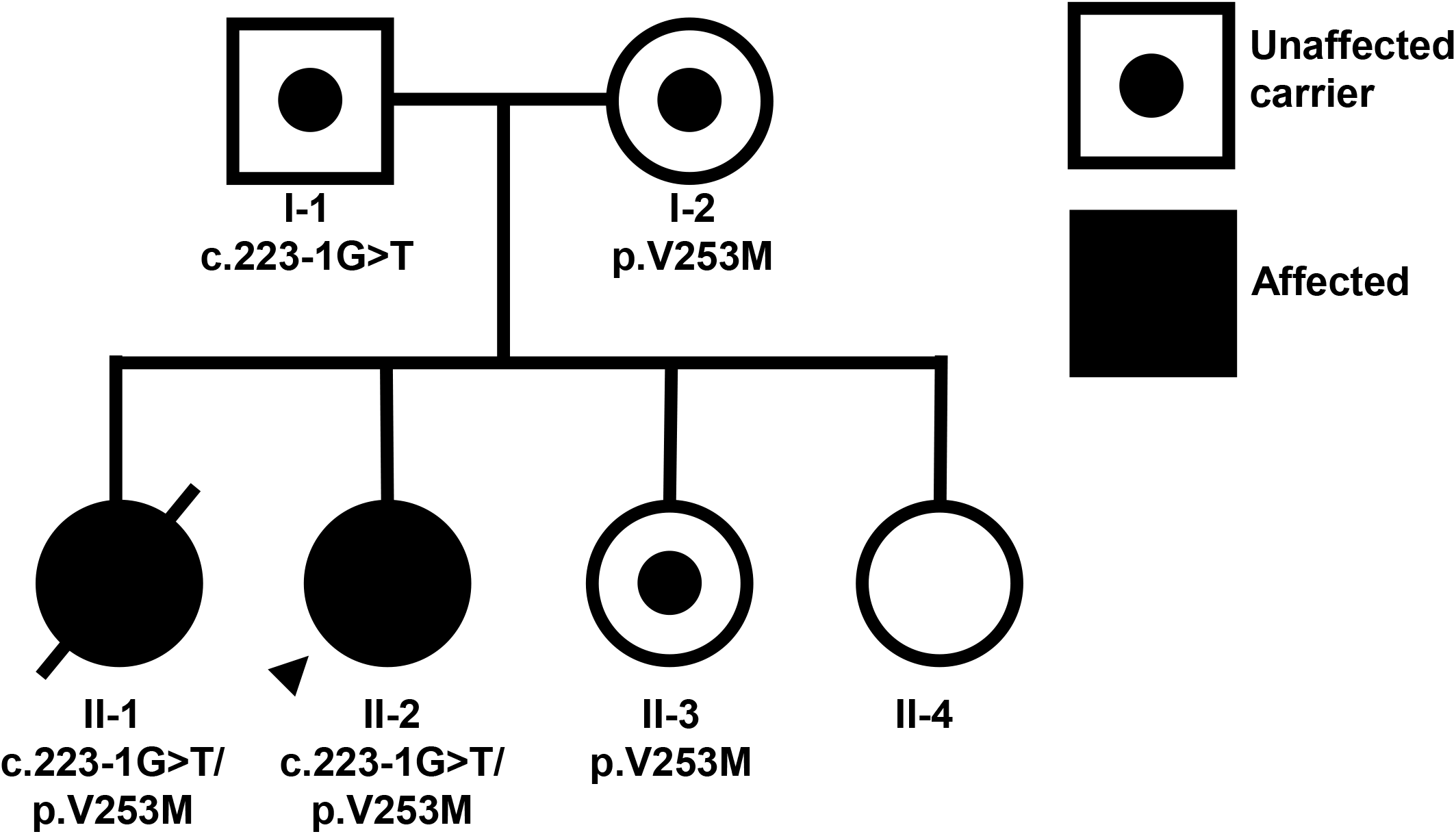
Family pedigree with paternal and maternal *KMO* variants.

### Exome analysis

Given that clinical evaluation did not reveal an underlying diagnosis, the proband was referred to the Baylor College of Medicine (BCM) Undiagnosed Diseases Network (UDN) for further evaluation. The exome data from the proband, her affected sister, and both parents were transferred for research analysis. No *de novo* variants were identified in either of the affected sisters. However, biallelic variants in *KMO* were noted in both affected sisters. The paternally inherited variant, NM_003679.4:c.223-1G>T, has an allele count of two with no individuals homozygous for the variant in gnomAD v4.1.1^29^ and impacts a canonical acceptor splice site in intron 3 of 14. The G nucleotide in the canonical splice site is conserved among rhesus macaques, mice, dogs, chickens, and zebrafish. SpliceAI predicts multiple aberrant splicing events including loss of the splice acceptor site (acceptor loss score of 0.95) of exon 4 and gain of an acceptor site 13 nucleotides downstream (acceptor gain score of 0.28)^30,31^. The maternally inherited variant, NM_003679.4:c.757G>A:p.(Val253Met), has an allele count of 158 in gnomAD 4.1.1 with no individuals homozygous for the variant, has a CADD score of 27.2^32,33^, and was predicted to be deleterious by multiple prediction programs including Sift^34^, Polyphen-2^35^, and Mutation Taster^36^. However, the AlphaMissense score for the maternally inherited variant is 0.204 (likely benign)^37,38^, and the REVEL score is 0.227^39^. The encoded valine is conserved across rhesus macaques, mice, dogs, chickens, *Xenopus*, and zebrafish. Based on annotations of KMO structures resolved in other organisms and a partially resolved human KMO crystal structure, this variant may be in a conserved domain seen in other flavoprotein monooxygenases^40–42^. Sanger sequencing demonstrated that one unaffected sister was heterozygous for only the c.757G>A variant, and the other unaffected sister had neither variant (**Figure 2**, II-3 and II-4, respectively). A search of gnomAD reveals only one homozygous predicted loss-of-function variant that impacts the last exon (exon 15) of *KMO*^29^.

RNA sequencing in whole blood was performed to confirm the impact of the c.223-1G>T variant on splicing. Although *KMO* expression is low in whole blood and, thus, a low number of reads were obtained, the findings supported the SpliceAI predictions of the creation of a new splice acceptor site 13 nucleotides into the exon in some reads (**Supplemental Figure 1**). This shift is expected to disrupt the reading frame leading to a frameshift and early stop codon. Although biallelic expression of *KMO* was noted, the low expression levels prevented strong conclusions about whether nonsense-mediated decay may be occurring at low or moderate levels.

### Kynurenine metabolites and NAD levels in the proband are altered in plasma and whole blood

*KMO* encodes kynurenine-3-monooxygenase, the enzyme that converts kynurenine to 3-hydroxykynurenine in the kynurenine pathway (**Figure 1**). To determine if the biallelic variants in *KMO* impact enzyme activity, we performed targeted metabolomics to assess kynurenine pathway metabolites upstream of KMO [tryptophan (TRP), kynurenine (KYN), kynurenic acid (KA), anthranilic acid (AA)], downstream of KMO [3-hydroxykynurenine (3HK), xanthurenic acid (XA), 3-hydroxyanthranilic acid (3HAA), quinolinic acid (QA)], Preiss-Handler and salvage pathway metabolites [nicotinic acid riboside (NAR), NAD^+^, nicotinamide mononucleotide (NMN), nicotinamide (NAM), nicotinamide riboside (NR)], and salvage pathway excretion products [1-methylnicotinamide (1MNA), N1-Methyl-2-pyridone-5-carboxamide (2PY), N1-methyl-4-pyridone-3-carboxamide (4PY)] in plasma from individuals I-1, I-2, II-2, and II-3. In plasma, compared with the mean of unaffected individuals, the proband (II-2) had a nearly 670% increase in KYN, the metabolite directly upstream of KMO, and a 2,153% increase in KA, which is synthesized from KYN by kynurenine aminotransferases (KATs)^43^. The proband had decreased levels of 3HK, XA, and QA and mildly lower NAD^+^ in plasma when compared to the unaffected mean (**Table 1**). In whole blood, only metabolites upstream and downstream of KMO were quantified using LC-MS/MS. The metabolites in whole blood from the proband demonstrated similar patterns as in plasma when compared to the unaffected mean, with an 833% increase in KYN, a 1594% increase in KA, and an approximately 63% decrease in XA (**Supplemental Table 1**).

**Table 1:** Levels of metabolites of the NAD metabolome are altered in plasma from family member II-2. Plasma levels of nicotinic acid (NA), nicotinic acid mononucleotide (NAMN), nicotinic acid adenine dinucleotide (NAAD), and nicotinamide adenine dinucleotide hydrate (NADH) levels were below the limit of quantification in all samples (data not shown). *Salvage pathway metabolites (particularly NAD^+^ and NMN) of individual I-2 may be affected by vitamin B3 supplementation, and thus, were excluded from calculations of unaffected mean and fold change in II-2.

| Metabolite | Individual and relation to proband |  |  |  |  |  | Mean in | Fold |
| --- | --- | --- | --- | --- | --- | --- | --- | --- |
|  | I-1,<br>Father | I-2,<br>Mother | II-2,<br>Proband | II-3,<br>Sibling | Control<br>female | Control<br>male | unaffected<br>individuals | change<br>in II-2 |
| **Kynurenine pathway, upstream of KMO** |  |  |  |  |  |  |  |  |
| TRP (μM) | 44.0 | 43.1 | 41.1 | 46.4 | 47.1 | 49.2 | 46.0 | 0.9 |
| KYN (μM) | 2.6 | 1.5 | 13.0 | 2.0 | 1.5 | 2.2 | 2.0 | 6.6 |
| KA (nM) | 222.9 | 100.4 | 2383.7 | 78.2 | 59.1 | 93.1 | 110.7 | 21.5 |
| AA (nM) | 26.8 | 24.5 | 246.7 | 30.7 | 21.3 | 23.5 | 25.3 | 9.7 |
| **Kynurenine pathway, downstream of KMO** |  |  |  |  |  |  |  |  |
| 3HK (nM) | 43.0 | 22.8 | 15.5 | 52.0 | 20.0 | 32.2 | 34.0 | 0.5 |
| XA (nM) | 37.8 | 25.0 | 16.4 | 18.8 | 21.4 | 33.5 | 27.3 | 0.6 |
| 3HAA (nM) | 59.9 | 34.0 | 17.4 | 31.7 | 25.0 | 37.0 | 37.5 | 0.5 |
| QA (nM) | 351.4 | 302.4 | 118.4 | 453.6 | 229.7 | 413.1 | 350.0 | 0.3 |
| **Preiss-Handler and salvage pathways** |  |  |  |  |  |  |  |  |
| NAR (nM) | 7.3 | 1.7 | 1.9 | 0.8 | 11.0 | 3.9 | 4.9 | 0.4 |
| NAD<sup>+</sup> (nM) | 4.4 | 49.6* | 1.4 | 2.4 | 9.2 | 7.2 | 5.8* | 0.2* |
| NMN (nM) | 0.7 | 4.3* | 1.3 | 0.9 | 3.3 | 3.2 | 2.0* | 0.7* |
| NAM (nM) | 73.0 | 92.1 | 73.7 | 141.6 | 70.4 | 85.7 | 92.6 | 0.8 |
| NR (nM) | 1.5 | 1.0 | 0.8 | 1.0 | 2.2 | 2.8 | 1.7 | 0.5 |
| **Excretion products** |  |  |  |  |  |  |  |  |
| 1MNA (nM) | 40.1 | 86.0 | 21.6 | 85.4 | 77.3 | 73.3 | 72.4 | 0.3 |
| 2PY (μM) | 1.1 | 1.4 | 0.6 | 1.2 | 2.8 | 1.6 | 1.6 | 0.4 |
| 4PY (nM) | 273.1 | 367.9 | 206.0 | 392.4 | 545.7 | 309.2 | 377.7 | 0.5 |

### *Kmo^-/-^* mice have a functional loss of KMO

The human metabolomic and splicing studies suggest that the variants are causing a loss of KMO function in the two affected sisters. Thus, we generated a *Kmo^-/-^* global knockout mouse model for additional mechanistic studies. CRISPR/Cas9 was used to delete exon 5 of 15 of *Kmo*. This deletion is predicted to cause a frameshift and early stop in exon 7, leading to nonsense-mediated mRNA decay (NMD) of *Kmo* and a functional decrease in KMO activity (**Supplemental Figure 2A**). To validate the deletion, we performed standard PCR on genomic DNA using primers surrounding exon 5, then Sanger sequenced the product and confirmed that the *Kmo^-/-^* mice have the expected deletion of exon 5 (**Supplemental Figure 2B**). In addition, *Kmo* expression in the liver of *Kmo^-/-^* mice was significantly decreased **(Figure 3A).** To confirm loss of KMO at the protein level, western blots were performed using adult kidney and liver tissue, which are two tissues with relatively high levels of *Kmo* expression^44^. Protein corresponding to KMO was not detected in samples from *Kmo^-/-^*mice (**Figure 3B**). As an antibody control, we used adult heart tissue from a *Kmo^+/+^* mouse, which is not expected to have *Kmo*/KMO expression. Furthermore, LC-HRMS demonstrated significantly increased levels of serum KYN in *Kmo^-/-^* mice, as expected with loss of KMO (**Figure 3C**).

**Figure 3.**
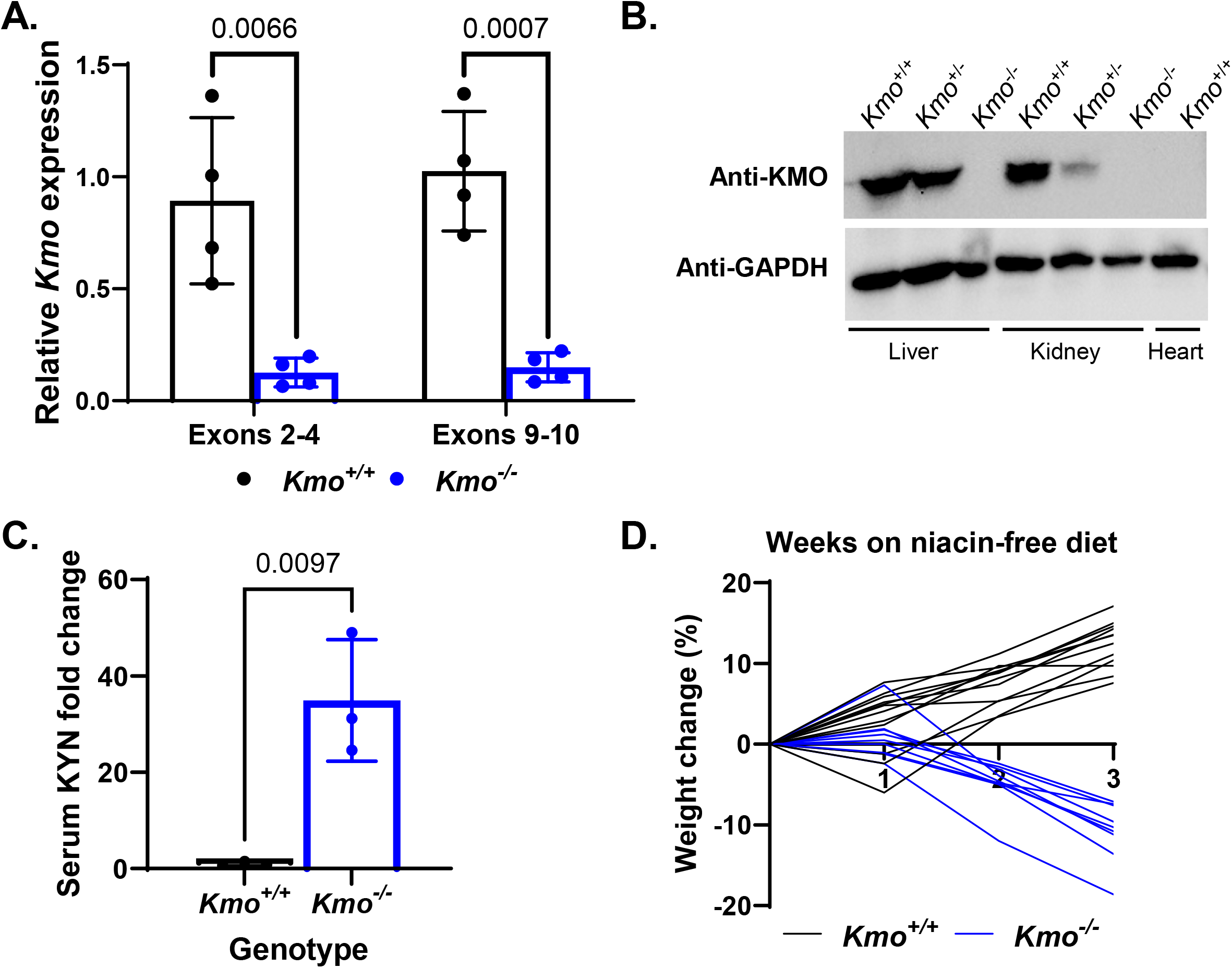
KMO mouse model validation. A) *Kmo* expression in adult mouse liver using primer pairs upstream and downstream of exon 5 (deleted exon). Welch’s unpaired t-tests, n = 4 each group. B) No band corresponding to KMO protein was observed in adult *Kmo^-/-^* mice through western blot using liver and kidney. Heart tissue from adult *Kmo^+/+^* animal lacks a KMO band, as expected. C) Serum kynurenine levels are increased in adult *Kmo^-/-^* mice compared to *Kmo^+/+^* mice. Raw data were normalized to the raw mean value of serum KYN from *Kmo^+/+^*mice, unpaired t-tests, n = 3 each group. D) *Kmo^-/-^* mice lose weight (up to 18%) over the course of three weeks on the NW0 diet, but their *Kmo^+/+^* littermates gain weight, demonstrating that KMO deficient mice are vitamin B3 dependent. n = 12 *Kmo^+/+^,* n = 9 *Kmo^-/-^*. A & C) Error bars represent mean and standard deviation (SD).

On a standard lab chow (5V5M in **Table 2**), the *Kmo^-/-^* mice are viable, fertile, and present at the expected Mendelian ratios similar to previously published *Kmo* knockout models^45–47^ (**Supplemental Table 2**). However, we hypothesized that *Kmo^-/-^*mice require dietary sources of vitamin B3 to synthesize NAD^+^ through the Preiss-Handler and/or salvage pathways and thus, would not survive on a vitamin B3-free diet. To test this hypothesis, we fed adult *Kmo^+/+^*and *Kmo^-/-^* mice a vitamin B3-free diet (NW0 diet in **Table 2**) for up to three weeks. As expected, *Kmo^-/-^* mice lost weight on the vitamin B3-free diet, whereas wildtype littermates on the same diet continued to gain weight (**Figure 3D**).

**Table 2:** Mouse diets and their approximate NAD precursors per day. NW refers to nicotinic acid in water. NAD precursors refer to nicotinic acid/niacin equivalents. According to LabDiet, the 5V5M diet has 0.26% tryptophan and 120ppm niacin. The standard diet (TD.97184, Teklad) is rich in nicotinic acid and tryptophan. The vitamin B3/nicotinic acid-free (NW0) diet (TD.220153, Teklad) is rich in tryptophan. *Calculations were made under the assumptions that 60 mg Trp is equivalent to 1 mg NA, the mice eat 3.9 g of food per day, and drink 6.2 mL water per day^24,74,75^.

|  | NA in feed<br>(mg/kg) | Tryptophan in feed<br>(mg/kg) | NA in water<br>(mg/L) | NAD Precursors*<br>(ug/day) |
| --- | --- | --- | --- | --- |
| **5V5M** | 120 | 2600 | 0 | 637 |
| **Standard diet** | 45 | 2000 | 0 | 305.5 |
| **NW0 diet** | 0 | 1800 | 0 | 117 |
| **NW8 diet** | 0 | 1800 | 8 | 166.6 |
| **NW16 diet** | 0 | 1800 | 16 | 216.2 |

### *Kmo^-/-^* embryos are at an increased risk for developing congenital anomalies when exposed to limited nicotinic acid during gestation

To assess whether *Kmo^-/-^* embryos are at increased risk for congenital anomalies in the setting of a diet low in vitamin B3, we generated *Kmo^-/-^* and *Kmo^+/-^* embryos from *Kmo^-/-^* dams fed a low nicotinic acid (NW8) diet vs. a control (NW16) diet (**Table 2**). Genotypes did not deviate from expected Mendelian ratios at E18.5 (**Supplemental Table 2**). However, at E18.5, approximately 50% of *Kmo^-/-^* embryos derived from dams fed the NW8 diet had ≥1 external anomaly detected through gross inspection by light microscopy and/or soft tissue anomaly detected through iodine-contrasted µCT as compared to *Kmo^+/-^* embryos (p=0.02, **Figure 4A**). However, there was no significant difference in the proportion of embryos with external and/or soft tissue anomalies when *Kmo^-/-^* and *Kmo^+/-^* embryos were derived from dams fed the NW16 diet (**Figure 4B**). Congenital anomalies detected through light microscopy included microphthalmia, anophthalmia, eyelid fusion, bent limbs, syndactyly, omphalocele, abnormal placenta, kinked/shortened tails, and other dysmorphic features. One *Kmo^+/-^* embryo was severely dysmorphic and was missing its lower mandible (**Supplemental Data 1, Sheet 1,** Sample ID: 5.4). Through iodine contrasted µCT, we were able to detect congenital anomalies with a higher sensitivity and identified embryos with cleft palate, complete renal agenesis, small kidneys, cardiac anomalies, vertebral/rib anomalies, and eye/optic nerve anomalies. Examples of anomalies are provided in **Figure 5** and **Supplemental Figure 3A and 3B**. A detailed list of anomalies detected per genotype and diet combination is provided in **Supplemental Data 1** and a bar graph showing percentages of embryos with ≥1 anomaly in a given region is shown in **Supplemental Figure 3C** (generated from data in Sheet 2 of **Supplemental Data 1**).

**Figure 4.**
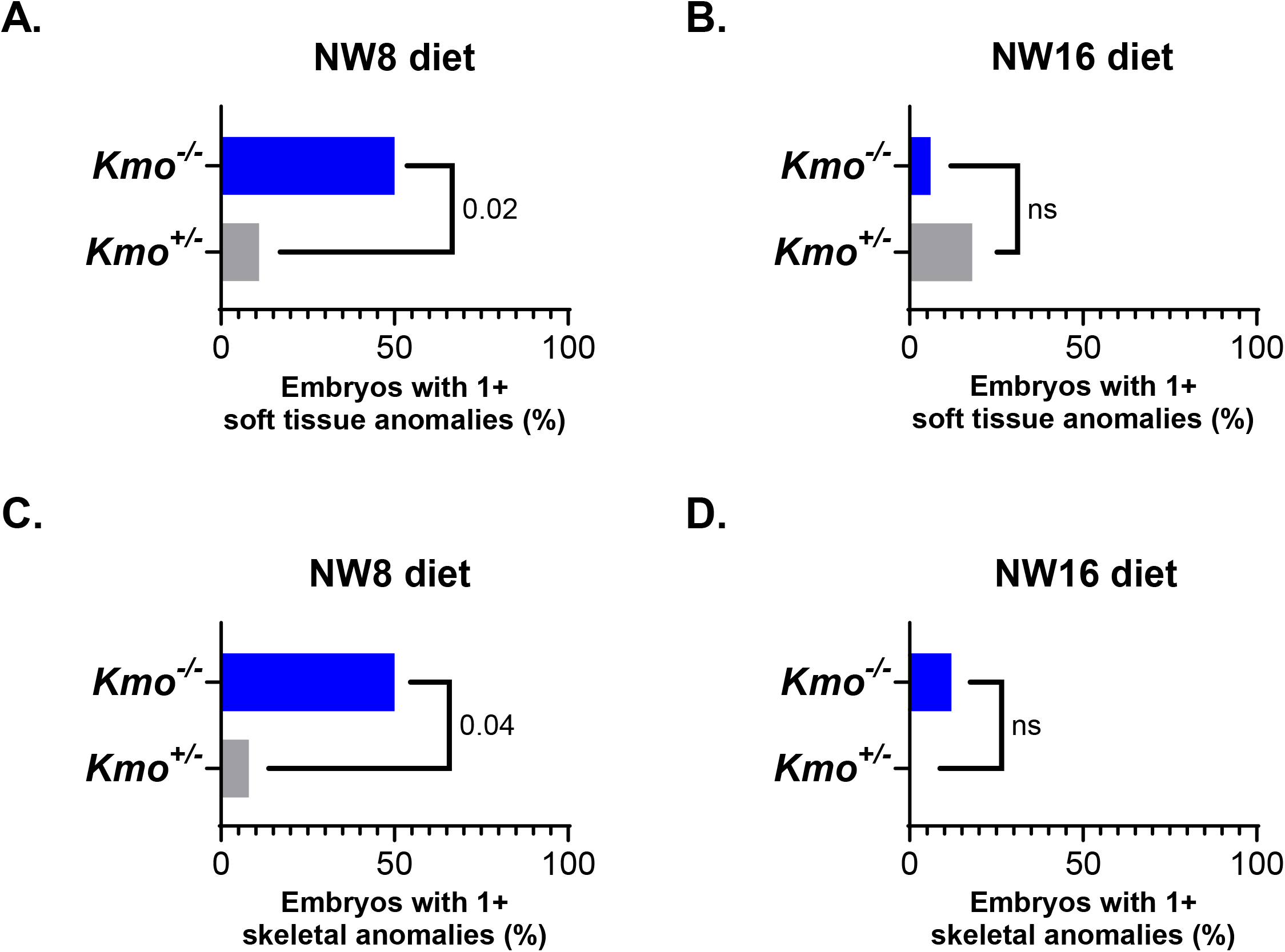
*Kmo^-/-^*embryos exposed to the NW8 diet are at a higher risk of developing congenital anomalies. A) *Kmo^-/-^* embryos derived from *Kmo^-/-^* dams fed the NW8 diet are at a higher risk of developing ≥1 soft tissue or external anomalies as compared to heterozygous littermates. n = 19 *Kmo^+/-^,* n = 16 *Kmo^-/-^*. B) No significant difference in soft tissue anomalies was detected in *Kmo^-/-^* and *Kmo^+/-^* embryos derived from *Kmo^-/-^* dams fed the NW16 diet. n = 17 *Kmo^+/-^,* n = 17 *Kmo^-/-^*. C) *Kmo^-/-^* embryos derived from *Kmo^-/-^* dams fed the NW8 diet are at a higher risk of developing ≥1 skeletal anomalies as compared to heterozygous littermates. n = 12 *Kmo^+/-^,* n = 14 *Kmo^-/-^*. D) No significant difference in the proportion of embryos with skeletal anomalies was detected embryos derived from *Kmo^-/-^* dams fed the NW16 diet. n = 17 *Kmo^+/-^,* n = 17 *Kmo^-/-^*. A-D) Two-tailed Fisher’s exact test. Comprehensive anomaly assessments and counts used to generate data in this figure can be found **Supplemental Data 1.**

**Figure 5.**
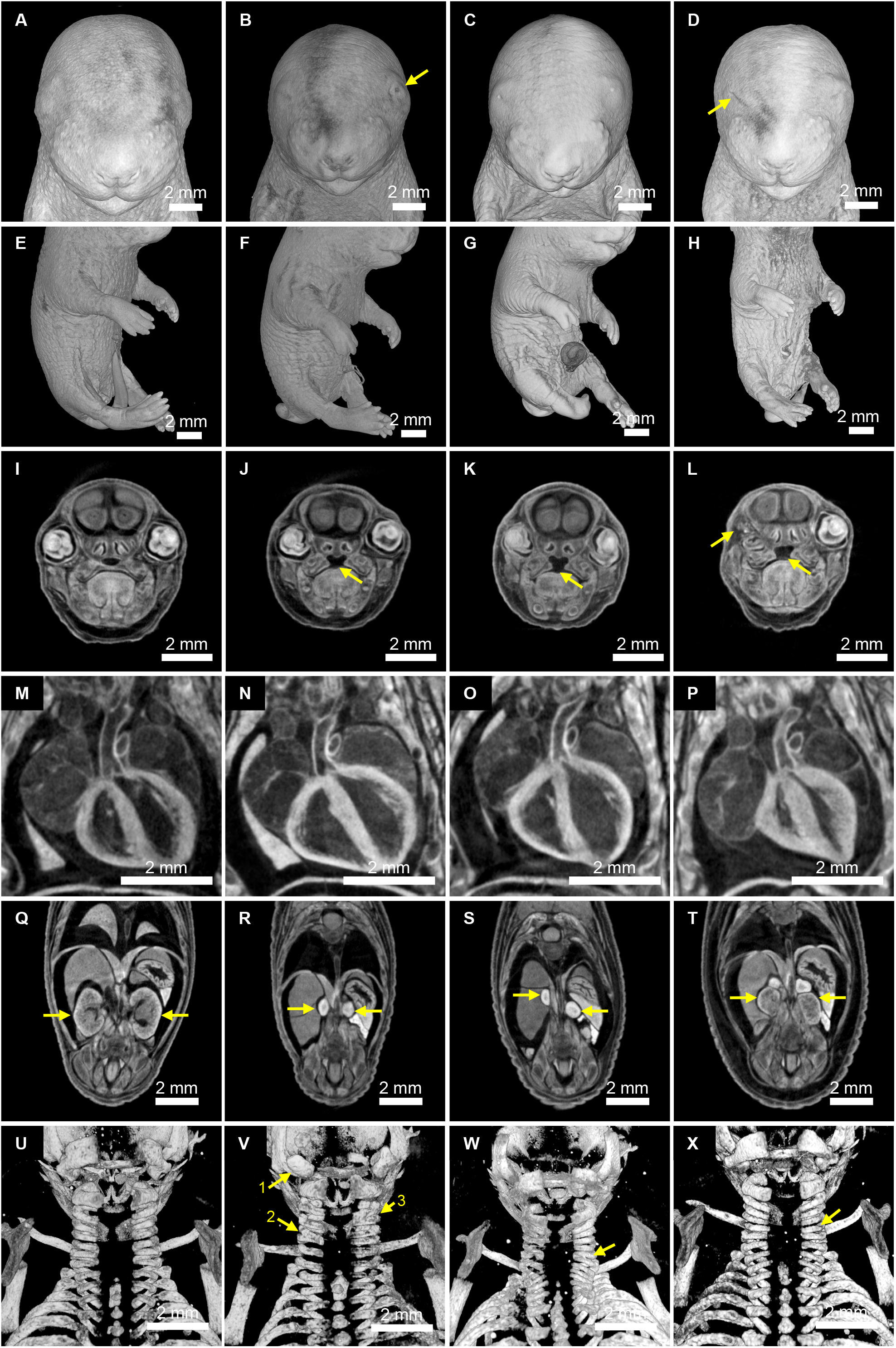
Examples of congenital anomalies in *Kmo^-/-^* embryos born to *Kmo^-/-^*dams fed the NW8 diet. Panels A, E, I, M, Q, and U are representative images of a morphologically normal *Kmo^+/-^* embryo (ID=6.3) exposed to the NW8 diet. B) Yellow arrow indicating eyelid fusion in a *Kmo^-/-^*embryo (ID=2.3). C) Bilateral microphthalmia in a *Kmo^-/-^* embryo (ID=2.5). D) Anophthalmia in a *Kmo^-/-^* embryo (ID=5.3). F, G, H) Kinked/curly tails in *Kmo^-/-^* embryos (left to right, IDs=2.3, 2.5, and 5.3). Omphalocele is also shown in panel G. J, K, L) Yellow arrows indicating cleft palate in *Kmo^-/-^* embryos (left to right, IDs=2.3, 2.5, and 5.3). Additional yellow arrow showing anophthalmia in panel L. N, O) Dilated cardiomyopathy in *Kmo^-/-^* embryos (left to right, IDs=3.4 and 3.9). P) Abnormal heart in a *Kmo^-/-^*embryo (ID=5.3). Q) Yellow arrows indicating normal kidneys in a *Kmo^+/-^*embryo (ID=6.3). R, S) Bilateral renal agenesis in *Kmo^-/-^* embryos with yellow arrows indicating adrenal glands with no kidneys beneath (IDs=5.3 and 2.3). T) Yellow arrows indicating small kidneys in a *Kmo^-/-^* embryo (ID=5.1). V) Vertebral anomalies in a *Kmo^-/-^* embryo (ID=2.3). Arrow 1 indicates abnormal C1, arrow 2 indicates a C5-C6 fusion, and arrow 3 indicates a C3-C4 fusion. W) Yellow arrow indicates a C7-T1 fusion in a *Kmo^-/-^*embryo (ID=4.1). X) Yellow arrow indicates a C6-C7 fusion in a *Kmo^-/-^* embryo (ID=4.8). Panels A-X) Embryo IDs listed in parentheses correspond to embryos that can be found in **Supplemental Data 1,** Sheet 1.

Because the two affected sisters also had vertebral anomalies, we performed µCT prior to administration of iodine contrast in a randomized subset of embryos to evaluate for skeletal abnormalities, such as vertebral and rib fusions or segmentation anomalies. At E18.5, *Kmo^-/-^* embryos from dams fed the NW8 diet had more cervical and thoracic spine fusion anomalies than *Kmo^+/-^* embryos (p=0.04, **Figure 4C**). On the NW16 diet, two *Kmo^-/-^*embryos developed a C5-6 fusion whereas none of the *Kmo^+/-^* littermates had skeletal phenotypes. Thus, there was no statistically significant difference in the proportion of *Kmo^-/-^*vs. *Kmo^+/-^* embryos with skeletal anomalies in the NW16 diet studies (**Figure 4D**).

Given the proband’s short stature, we also compared the crown-rump length of the *Kmo^-/-^* vs. *Kmo^+/-^* embryos on both diets. The *Kmo^-/-^* embryos had significantly reduced crown-rump length as compared to *Kmo^+/-^* embryos exposed to the NW8 diet *in utero* (p-value=0.04, **Supplemental Figure 4A**). In contrast, there was no significant difference in the crown-rump length in *Kmo^-/-^* and *Kmo^+/-^* embryos exposed to the NW16 diet (p-value=0.46, **Supplemental Figure 4B**).

To further analyze this difference in size, we compared the mineralized long bone lengths in the *Kmo^-/-^* vs. *Kmo^+/-^* embryos on both diets. This analysis revealed significantly shorter ulna lengths in *Kmo^-/-^* embryos as compared to *Kmo^+/-^* embryos exposed to the NW8 diet *in utero* (p-value=0.0007, **Supplemental Figure 5A**). However, the femurs and radii only trended towards significance in the same group (p-value=0.08 and p-value=0.06, respectively, **Supplemental Figures 5C and 5E**). *Kmo^-/-^* and *Kmo^+/-^* embryos exposed to the NW16 diet *in utero* did not have any differences in bone lengths (p-values>0.4, **Supplemental Figures 5B, 5D, and 5F**).

### Exploratory embryo metabolomics reveal alterations in pyrimidine, purine, and pentose phosphate metabolism in *Kmo*-deficient embryos

E11.5-E12.5 represents a critical timepoint for cardiac and renal development in mice^10,12,24,48,49^, so we hypothesized that NAD measurements at this stage could indicate whether organogenesis is occurring in the setting of low NAD in *Kmo^-/-^* embryos exposed to the NW8 diet *in utero*. To test this hypothesis, we measured NAD^+^ levels in whole embryos collected at E12.5. At this timepoint, genotypes did not deviate from expected Mendelian ratios (**Supplemental Table 2**). Moreover, the E12.5 embryos had similar weights regardless of genotype and dietary exposure (**Supplemental Figure 4C and 4D**). However, *Kmo^-/-^* embryos from dams fed the NW8 diet at E12.5 had significantly lower whole-body NAD^+^ levels than their littermates (p-value=0.01, **Figure 6A)**. In contrast, *Kmo^-/-^* embryos exposed to the NW16 diet had comparable NAD^+^ levels to their heterozygous littermates (**Figure 6B**).

**Figure 6.**
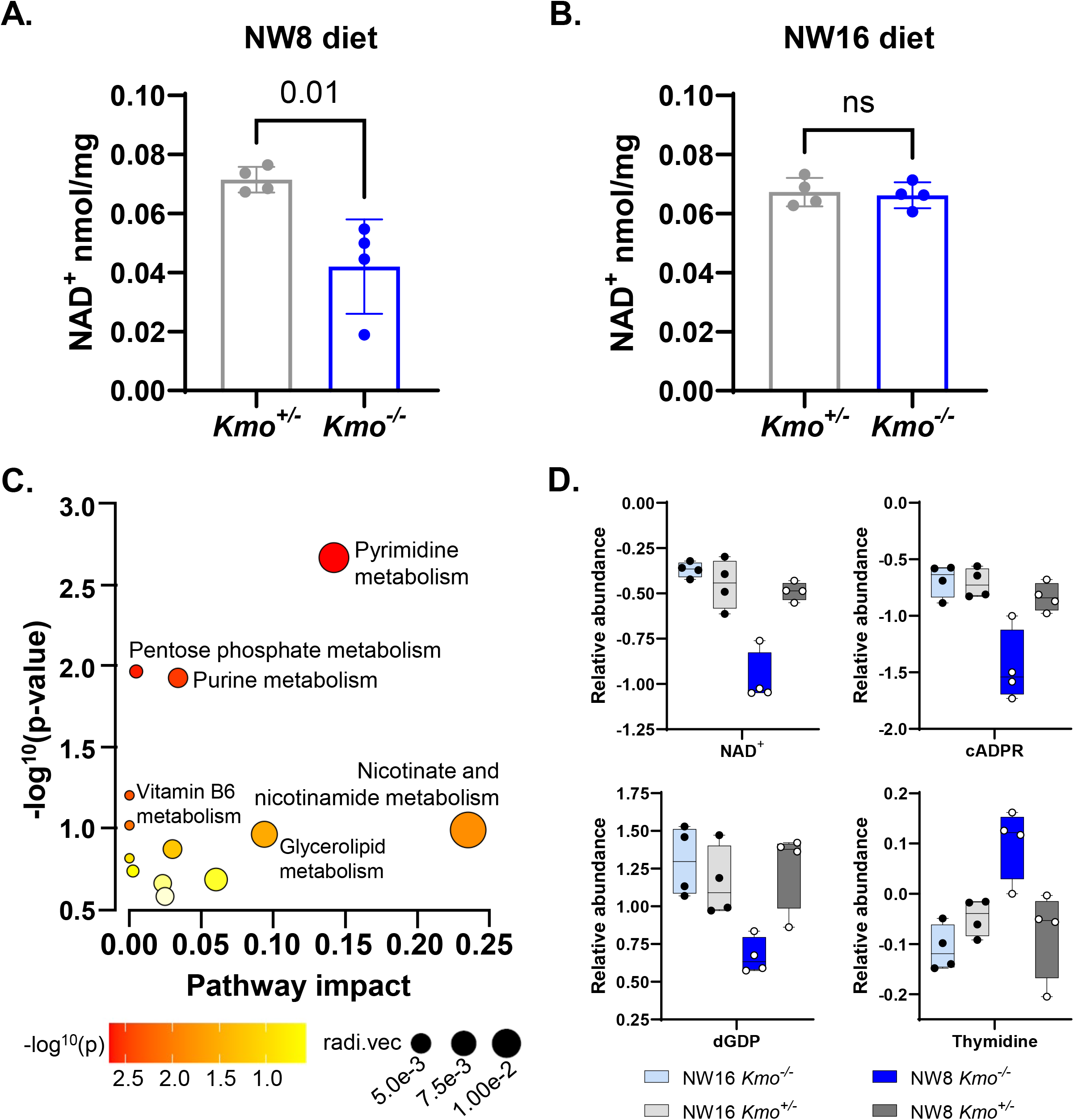
Murine embryo metabolomics demonstrate changes in NAD synthesis and purine and pyrimidine metabolism. A-B) Only *Kmo^-/-^* embryos derived from *Kmo^-/-^* dams fed the NW8 diet have low NAD^+^ levels (E12.5). Unpaired t-tests, n = 4 each group, error bars represent mean and SD. C) Pathway analysis of murine embryo features/metabolites detected through untargeted metabolomics that were significant for interaction effects (FDR ≤0.25). Using MetaboAnalyst 6.0^50^, pathways with metabolites that were enriched in the analysis included pyrimidine, pentose phosphate, purine, and nicotinamide/nicotinate pathways. radi.vec is a visualization of the impact score and was calculated internally by MetaboAnalyst using the impact score of the metabolite and pathway. Metabolites were mapped to *Mus musculus* KEGG pathways. D) Examples of metabolites that were significant for interaction effects that were mapped to KEGG pathways by MetaboAnalyst and that drive the results seen in panel C. Error bars represent maximum and minimum values. This is a subset of the data provided in **Supplemental Figure 9** and **extended Supplemental Figure 9**.

To explore metabolic perturbations that are associated with a *Kmo* deficiency, we performed exploratory untargeted metabolomics in E12.5 *Kmo^-/-^* and *Kmo^+/-^* embryos exposed to either the NW8 diet or the NW16 diet (**Figure 6C**). Using a two-way ANOVA with an FDR threshold of ≤0.25 and p-value<0.05, we identified metabolites that exhibited significant effects for genotype, diet, or genotype × diet interaction in the four groups of embryos (**Supplemental Figures 6-9, Supplemental Data 2**). A Tukey’s post-hoc for multiple comparisons was run on the metabolites with a significant genotype × diet interaction to determine which metabolites were uniquely high or low in the *Kmo^-/-^* embryos exposed to the NW8 diet as compared to the other three groups of embryos. First, as expected, the untargeted metabolomics confirmed a reduction in NAD^+^ and in cyclic ADP-ribose (cADPR), which is synthesized from NAD^+^, in the *Kmo^-/-^* embryos exposed to the NW8 diet. Interestingly, there were also several metabolites in the purine degradation pathway that were increased in the *Kmo^-/-^* embryos exposed to the NW8 diet including inosine and hypoxanthine, although it was not unique to the group via Tukey’s post-hoc tests (**Supplemental Figure 9, extended** and **Supplemental Data 2**). N6-succinyladenosine appears to be higher in the *Kmo^-/-^* embryos exposed to the NW8 diet but was only significant through a diet effect. Similarly, the pyrimidines, thymine and thymidine, were increased whereas the purine, deoxyguanosine diphosphate (dGDP), was decreased in the NW8 *Kmo^-/-^*embryos.

To further explore potential pathway enrichments, we performed a pathway analysis with metabolites that had FDR ≤0.25 for significant interaction effects (MetaboAnalyst 6.0^50^). This analysis revealed enrichments for the pyrimidine, pentose phosphate, and purine pathways (**Figure 6C and 6D**). Altogether, these results indicate that NAD deficiency may compromise energy metabolism, with downstream effects on purine and pyrimidine metabolic pathways during development.

## Discussion

The phenotypes observed in the two affected sisters reported in our study overlap with the phenotypic spectrum reported in CNDD, which typically manifests with the co-occurrence of cardiac, renal, vertebral, and limb anomalies^10–22^. Biallelic variants in *KYNU, HAAO,* and *NADSYN1* associated with CNDD have heterogeneous clinical manifestations, ranging from perinatal lethality and congenital anomalies incompatible with life to comparatively mild skeletal abnormalities with survival into adulthood^10–16,20,21^. Both affected sisters with biallelic *KMO* variants had severe congenital heart anomalies, vertebral anomalies, and short stature, while the surviving child also had small kidneys, unilateral hearing loss, and learning disabilities. In addition to congenital anomalies, the biochemical findings in the proband further support a diagnosis of CNDD. Targeted metabolite analyses in the proband demonstrate strongly elevated levels of the intermediates of NAD *de novo* synthesis upstream of KMO (KYN, KA, AA) and lower levels of those downstream (3HK, XA, 3HAA, QA), as well as moderately lower NAD^+^ and NAD excretion metabolite levels (**Table 1, Supplemental Table 1**). In contrast, plasma NAD^+^ is not always low in individuals with CNDD and thus may not be a reliable marker for the disorder^12^. Apart from elevated maternal NAD^+^ and NMN levels, the unaffected family members had NAD metabolite levels similar to those reported for healthy non-pregnant women^51^, indicating their NAD metabolism was not affected. These data establish KMO deficiency as a novel form of CNDD and expand the genetic heterogeneity of this disorder.

Given the nature of the anomalies and the hypothesized gene × environment interaction, we used a mouse model to test our hypothesis that KMO deficiency increases the risk for congenital anomalies in the setting of a maternal diet low in NAD precursors, such as nicotinic acid. There are fundamental differences between NAD metabolism in humans and mice that impacted the design of our mouse studies. In mice, tryptophan is efficiently converted to NAD using the kynurenine pathway^10,12,23,24,52,53^. In humans, there is an increased shuttling of tryptophan metabolites away from NAD synthesis and towards the TCA cycle (**Figure 1**)^53,54^. As a result, and because mice have a higher food consumption and metabolic rate relative to their body size, humans are more susceptible to insufficient availability of usable NAD precursors as compared to mice. This is evidenced by the observation that *Kmo^-/-^* mice are viable, fertile, and have no apparent congenital anomalies on standard chow^45,47,55–57^, whereas the proband’s mother is heterozygous for the *KMO* variant, yet she had CNDD-affected children (II-1 and II-2). The *Kmo^-/-^* mice in our study failed to thrive on a vitamin B3-free diet, demonstrating a functional dependence on vitamin B3-derived NAD precursors. Therefore, to better model the stronger dependence on sufficient usable NAD-precursor provision in humans, and specifically pregnant women, we used *Kmo^-/-^* (dam) × *Kmo^+/-^* (stud) crosses rather than heterozygous intercrosses to generate embryos and controlled the NAD precursor intake via the water and chow, similarly to previously published studies of other mouse models of CNDD genes^10,12,23,24^. Our embryo studies support a gene × environment interaction like those described in other forms of CNDD. *Kmo^-/-^* embryos exposed to the low nicotinic acid diet (NW8) had a significantly higher incidence of congenital anomalies as compared to their heterozygous littermates or embryos exposed to the control diet (NW16). The spectrum of congenital anomalies was similar to those seen in previous studies with *Kynu*^-/-^ and *Haao*^-/-^ CNDD mouse models on a slightly more NAD precursor-restricted diet providing 159.7 µg of NAD precursors per day^10,24^. The affected *Kmo^-/-^* embryos also developed phenotypes that overlap with those seen in the affected sisters, including vertebral anomalies, cardiac defects (including DORV in one embryo, **Supplemental Figure 3B**), renal abnormalities, and reduced growth.

Prior studies of CNDD have established that impaired NAD synthesis is associated with congenital anomalies and pregnancy loss. Consistent with these studies, the proband had low NAD^+^ levels, and *Kmo^-/-^*embryos exposed to the low vitamin B3 diet had significantly reduced NAD^+^ levels during organogenesis. However, the targeted metabolomics in our proband showed extreme elevations in KYN and KA that exceed those reported in other CNDDs (**Table 1, Supplemental Table 1**). Thus, KMO deficiency causes extreme elevations of the upstream metabolites, KYN and KA, as compared to 11 previously published cases of CNDD/VCRL with targeted or untargeted metabolomic profiles^10,12,15,58^. Although elevations in KYN and KA are associated with inflammation/immune phenotypes and neuropsychiatric disorders, the consequences of such extreme elevations of these metabolites in KMO deficiency are unclear. Altogether, these metabolite findings demonstrate that clinically available metabolic profiling can detect disruptions in NAD synthesis pathways in CNDD and may serve as a tool for evaluating the pathogenicity of variants of uncertain significance in *KMO* and other kynurenine pathway genes^12^. Moreover, the human and adult mouse metabolite data support elevated KYN and KA as the most suitable biomarkers for KMO deficiency.

Both targeted NAD^+^ analyses and exploratory untargeted metabolomics in E12.5 murine embryos confirmed that only *Kmo^-/-^* embryos exposed to the NW8 diet had significantly lower NAD^+^ levels. Organogenesis is a highly energy-demanding period of development^10,12,24,48,49^ and is therefore particularly vulnerable to energy failure resulting from NAD deficiency. Metabolomic patterns in *Kmo^-/-^* embryos fed the NW8 diet suggest that this energy deficit may be associated with disruptions in purine and pyrimidine metabolism (**Supplemental Data 2**). The changes in purine and pyrimidine metabolism may impair DNA/RNA synthesis and DNA repair, contributing to pregnancy loss and congenital anomalies. Alternatively, reduced levels of cADPR—an NAD⁺-derived second messenger that regulates Ca²⁺ release—may disrupt cell signaling during this critical developmental window and contribute to the anomalies observed in this disorder. Although we were able to measure cADPR, adenosine, and ADP in the murine embryos, the platform we used did not detect adenosine diphosphate ribose (ADPR). Altered ADPR levels can indicate the consumption of NAD^+^ by NAD-dependent enzymes, such as Poly ADP-ribose polymerases (PARPs). Thus, a third possibility may be that PARPs, which are necessary for DNA damage repair or protein-protein interactions, are unable to function properly due to low NAD. However, it is interesting to note that metabolites within the kynurenine pathway, such as KYN and KA, which were elevated in the serum of adult *Kmo^-/-^* mice and the proband’s samples, were not elevated in *Kmo^-/-^* embryos exposed to the NW8 diet (comparable across the four groups, see **Supplemental Data 2** and raw data available upon request). This may be due to a maternal clearance of these metabolites or a compartmentalization effect due to the differences in sample types (e.g., whole embryo vs. plasma from the proband).

The phenotypic variability among the *Kmo^-/-^* embryos exposed to the NW8 diet raises many questions about the nature of this gene × environment interaction. A previous study of CNDD in mice suggests that maternal NAD availability as measured in whole blood influences the variability of congenital anomalies in NAD-deficient embryos^52^. Thus, the phenotypic heterogeneity may reflect differences in NAD^+^ availability secondary to maternal factors such as variability in maternal water consumption. It is unclear whether this phenotypic variability may also be an explanation for why we have only identified one family and one living individual with CNDD caused by a KMO deficiency. The most severe end of the NAD^+^-deficient mouse embryonic phenotype is resorption^12,23,24,52^. Thus, complete KMO deficiency may lead to pregnancy loss in humans. Because the c.223-1G>T variant impacting a canonical splice acceptor site is expected to cause NMD, perhaps the p.Val253Met variant leads to residual KMO activity, which enables survival. Alternatively, sufficient vitamin B3 intake during pregnancy might mask the phenotype. However, if this was the case, we would expect to find individuals with homozygous loss-of-function variants in databases, such as gnomAD, with healthy individuals, yet a search of gnomAD^29^ did not identify any predicted loss of function of variants at positions that would be predicted to result in NMD.

Together, our studies establish KMO deficiency as a novel cause of CNDD. By integrating metabolomic profiling from the proband with *Kmo^-/-^* mouse model studies, we demonstrate that loss of KMO disrupts NAD *de novo* metabolism in both mice and humans. In our proband and adult mice, KMO deficiency causes extreme elevations of upstream metabolites such as KYN, KA, and/or AA. In our murine embryos, loss of KMO is associated with lower NAD⁺ levels, and appears to perturb several metabolic pathways, including purine and pyrimidine metabolism, only in the setting of a maternal diet low in vitamin B3. Our findings support a model in which insufficient NAD availability may impair nucleotide metabolism or other NAD-dependent processes during key developmental windows, potentially increasing the risk for structural birth defects. These findings highlight a key gene × environment interaction that is critical for human development. Future studies using tissue-specific knockouts and in-depth metabolic and transcriptional analyses will be essential to define the precise pathogenic mechanisms and to determine whether prenatal detection or targeted NAD-precursor supplementation may be beneficial to patients with this diagnosis.

## Methods and Materials

### Sex as a biological variable

Male and female mice were used in this study. Sex was not considered as a biological variable. *Sry* genotyping to determine murine embryo sex was performed using primers Sry F and Sry R listed in **Supplemental Table 3** and standard polymerase chain reaction (PCR) with an annealing temperature of 58°C. Sex of E18.5 embryos is recorded and stated in **Supplemental Data 1.** For untargeted metabolomics, two male and two female E12.5 embryos were included in each of the four groups.

### Trio exome sequencing

Clinical trio exome sequencing was performed at Baylor Genetics as described previously^59^ and analyzed with the proband exome data previously generated for the proband’s deceased sister. Exome data from both parents and both affected sisters were transferred to the UDN for research analysis. For the research analysis, all biallelic variants that were shared between the proband and her deceased sister with a minor allele frequency of <0.01% in the general population (gnomAD v2.1.1^60^) and in our internal database were prioritized for review. Variants observed as homozygous in multiple individuals in gnomAD were not prioritized for analysis. Variants of interest were confirmed by Sanger sequencing.

### Human metabolomic studies

Fasted whole blood and plasma samples from family members I-1, I-2, II-2, and II-3 were collected, stored at -80°C, and used for targeted metabolomics. Targeted analysis of NAD^+^ and related metabolites was performed in plasma and whole blood, as previously described, using liquid-chromatography tandem mass spectrometry (LC-MS/MS)^10,51^. Individual I-2’s NAD^+^ and NMN values may be affected by vitamin B3 supplementation, so they were excluded from calculations for the unaffected mean and fold change in II-2.

### RNA sequencing

Blood samples were collected from the proband and stored in PAXgene tubes at -80° C prior to extracting RNA. RNA extraction^61^, processing, sequencing, and analysis were performed as previously described^62^. SnapGene® software (version 8.0.2, from Dotmatics) was used to analyze the splice products that were identified using RNA sequencing.

### Animals

Mice used in this study were housed in microisolator cages in ventilated racks in an AAALAC accredited animal facility at BCM. The rooms were temperature and humidity controlled with a light:dark cycle of 6:00 AM to 8:00 PM. For regular maintenance of the colony, the mice were given *ad libitum* access to food (PicoLab® Select Rodent Diet 50 IF/9F Diet (Lab Diet, Cat# 5V5R), unless otherwise specified) and autoclaved drinking water. All studies were approved by the BCM Institutional Animal Care and Use Committee.

### *Kmo^-/-^* mouse model generation

We generated a *Kmo* global knockout (*Kmo^-/-^*) mouse model using CRISPR/Cas9 to generate an interval deletion of exon 5 which was predicted to produce a frameshift and premature termination. To do this, sgRNA sequences were designed to flank the critical exon 5 using the following guides: 5’GTGTGACGGTAACATGGGTTggg (https://wge.stemcell.sanger.ac.uk/crispr/317296880), and 5’CTTAGAGGTTACAGCCTCGAggg (https://wge.stemcell.sanger.ac.uk/crispr/317296939). The sgRNAs were selected by the BCM Genetically Engineered Rodent Model (BCM GERM) Core using the Wellcome Sanger Institute Genome Editing (WGE) website^63^. The BCM GERM Core electroporated a mixture consisting of precomplexed sgRNA (6.6 μM total gRNA concentration) and Cas9 Protein (6 μM, PNA Bio) in a final volume of 10 μL 1xTE (RNAse-free) into at least 60 pronuclear stage C57BL/6J (Jackson Lab Strain #:000664; RRID:IMSR_JAX:000664) zygotes as previously described^64^. Founder mice harboring an exon 5 deletion were identified using standard PCR with primers designed to amplify across the deletion interval (58°C annealing temperature, Kmo K1 F and Kmo K1 R, **Supplemental Table 3**). A second pair of primers was used to identify the endogenous allele, with one primer residing in the predicted deletion region (Kmo E1 F and Kmo E1 R, **Supplemental Table 3**). The expected size for the endogenous (wild type) PCR product is 460 bp, and the expected size for the deletion (knockout) product is ∼460 bp. In a wild-type allele, the knockout primer pair produces a product that is too large to be amplified using routine PCR protocols. F0 mice identified with the deletion interval were mated to stock, wild type C57BL/6J mice, and N1 progeny were screened using the same strategy to identify heterozygous knockout (*Kmo^+/-^*) animals. The deletion was confirmed using Sanger sequencing of PCR products amplifying across the deletion interval (**Supplemental Figure 4**). Sequence verified *Kmo^+/-^* N1 mice were backcrossed to C57BL/6J mice to generate N2 mice. The line was maintained as an intercross.

### Quantitative real time PCR

Quantitative real time (RT)-PCR was performed using RNA isolated from liver tissue from *Kmo^-/-^*and *Kmo^+/+^* mice. Upon dissection, tissues were snap frozen in liquid nitrogen and stored at - 80°C. Total RNA was extracted using a kit following manufacturer instructions (Qiagen RNeasy Mini Kit, #74104, and RNase-Free DNase Set, #79254). RNA purity and quality were assessed with a Nanodrop (Thermo Scientific, Nanodrop 2000c Spectrophotometer) and by verifying the presence of bands corresponding to 18S and 28S ribosomal RNA on a 1.5% agarose gel. cDNA was synthesized from RNA using the SuperScript III First-Strand Synthesis System for RT-PCR (Invitrogen, #18080-051). RT-qPCR was performed using a Quantstudio 7 Flex (Applied Biosystems) in triplicate using PowerUp SYBR Green Master Mix for qPCR (Applied Biosystems, #A25742). RNA expression of *Kmo* in liver was assessed with primers upstream of the deleted exon flanking exons 2-3 (Kmo E2-4 F) and exons 3-4 (Kmo E2-4 R) as well as primers downstream of the deletion in exon 9 (Kmo E9-10 F) and exon 10 (Kmo E9-10 R). Mouse ꞵ-actin was used as the reference gene (Bact-F and Bact-R); see **Supplemental Table 3** for primer sequences. Fold change in gene expression was calculated using the 2−ΔΔCt method^65^.

### Western blotting

To further verify loss of KMO protein, western blotting was performed using heart, liver, and kidney tissues from adult *Kmo^-/-^*, *Kmo^+/-^*, and *Kmo^+/+^* mice. Tissues were collected from adult mice that were euthanized via isoflurane inhalation. Tissues were cut into 50 mg pieces, snap frozen in liquid nitrogen at time of dissection, and stored at -80°C until further processing. RIPA buffer (Thermo Scientific, #89900) containing protease and phosphatase inhibitors (Roche, #4693159001 and #4906845001) was added to each 50 mg sample, then a tissue lyser (TissueLyser II, Qiagen) was used to homogenize the tissue (1 minute × 3, vibration frequency 26). The samples were centrifuged for 10 minutes at 13,000 rpm at 4°C, the supernatant was collected, and protein concentrations were measured using the Pierce BCA protein assay (Thermo Scientific, #23227). Laemmli buffer (Bio-Rad, #1610747) and β-mercaptoethanol were added to each sample then boiled for five minutes at 99°C. The samples were then run on a 10% mini protean TGX precast gel (Bio-Rad, #456-1084) and transferred to PVDF membranes (Millipore Sigma, #IPFL00010). Antibodies used for detection included a monoclonal anti-KMO (1:1000, R&D Biotechne, #MAB8050-SP), anti-rabbit IgG (1:10,000, Sigma-Aldrich, #A0545), and anti-GAPDH antibody (1:10,000, Sigma-Aldrich, #G9295). According to the R&D Systems datasheet, the anti-KMO antibody was raised against an immunogen that contained recombinant human KMO amino acids Asp2-Leu441 (UniProt #O15229), which contains the sequence for exon 5 of *KMO*.

### Genotyping

To genotype embryos, DNA was extracted from yolk sacs for PCR-based genotyping. To genotype adult mice, DNA was extracted from ear or tail biopsies. PCR was performed with amfiSure PCR Master Mix(2X) (GenDEPOT, #P0311-125) using the same two-primer system used to identify founder animals (**Supplemental Table 3,** Kmo E1 F and Kmo E1 R; Kmo K1 F and Kmo K1 R).

### Validation of functional deficiency of KMO in *Kmo^-/-^* adult mice

To confirm functional KMO deficiency in *Kmo^-/-^* mice, we collected serum from n=3 *Kmo^-/-^* and n=3 *Kmo^+/+^*mice aged 5-7 weeks that were fed Lab Diet 5V5R (**Table 2**). Mice were fasted for 3 hours prior to sample collection, anesthetized via isoflurane inhalation, then a cardiac puncture was performed to collect blood. Blood was placed into a yellow-top serum separator tube and incubated at room temperature for 30 minutes. The tubes were centrifuged for 5 minutes at 5,000 rpm at room temperature to separate the serum, then aliquoted into a 1.5mL Eppendorf tube and snap frozen. Serum was stored at -80°C until metabolite measurements.

To determine whether *Kmo^-/-^* mice were vitamin B3-dependent, *Kmo^+/-^* mice were intercrossed, and their pups were weaned onto Lab Diet 5V5R at 3 weeks of age and maintained on the diet until aged 6-7 weeks. Starting at 6-7 weeks of age, male and female *Kmo^-/-^* and *Kmo^+/+^* mice were weighed and placed onto vitamin B3-free chow (TD.220153, Teklad) without added nicotinic acid in the water (NW0 diet, **Table 2**). Body weight was recorded weekly for three weeks.

### Diet studies and phenotyping in *Kmo^-/-^* and *Kmo^+/-^* embryos

Prior to timed matings, female *Kmo^-/-^* mice and male *Kmo^+/-^* mice were fed a “standard” defined diet that was irradiated prior to delivery (TD.97184, Teklad; lot numbers: 3416722, 3468451, 3526761, 3562404, 3604343, 3649227, 3777472, 3814211, 3833075, 3911782) for at least 3 weeks. This diet contained tryptophan and nicotinic acid as the only source of vitamin B3 (Standard diet, **Table 2**). For embryo studies, timed matings were set up between male and female mice during the afternoon. When a vaginal plug was confirmed the next morning, female and male mice were separated, and embryos were considered embryonic day (E) 0.5. Starting at E0.5, dams were singly housed and fed a vitamin B3-free chow (diet was irradiated before delivery, TD.220153, Teklad; lot numbers: 3433936, 3526841, 3602091, 3760401, 3814202, 3833067, 3911791) and autoclaved drinking water that contained defined concentrations of United States Pharmacopeia (USP)-grade nicotinic acid (Spectrum Chemical Mfg Corp, #NI100, lot numbers: 1KB0557, 1LF0456, and 1MG0073) (NW8 diet and NW16 diet, **Table 2**). Water containing nicotinic acid was made fresh each week. Chow was replaced at least once per week. Pregnant mice were maintained on these diets until embryo collections at E12.5 for NAD^+^ measurements and untargeted metabolomics or at E18.5 for embryo phenotyping.

For embryos collected at E12.5, dams were euthanized via CO_2_ inhalation. Once euthanasia was confirmed, embryos were harvested, weighed, and snap frozen in liquid nitrogen. E12.5 embryos were then stored at -80°C until further processing. For embryos collected at E18.5, dams were euthanized via prolonged CO_2_ inhalation for 1 hour and dissected after confirmation of euthanasia^66,67^. E18.5 embryos were subsequently analyzed with light microscopy blinded to genotype and treatment group. For μCT imaging with and without iodine contrast, after E18.5 embryos were collected, they were fixed in 4% paraformaldehyde (PFA) and stored at 4°C until processing for μCT imaging. Prior to imaging, the embryos were embedded in 1% weight/volume (w/v) agarose, as previously described^66,67^. The Skyscan 1272 micro-CT scanner (Bruker) was used to acquire raw data for 3D imaging of the samples. The projection images thus obtained were reconstructed using the NRecon Reconstruction software (Version 1.6.9.8; Bruker). The reconstructed 3D data was rendered for volumetric visualization utilizing CTVox (Version 2.6.0; Bruker) or alternatively, Slicer freeware (Version 5.5.2; www.slicer.org)^68^. Annotation of soft tissue and skeletal anomalies was performed in a blinded manner on each embryo by three independent researchers. One researcher performed embryo bone measurements. Due to high rates of hydronephrosis in mice on the C57BL/6 background^69^ and in an embryo control group (21.87%, 14/64 embryos), we excluded these phenotypes from our anomaly counts. Due to high rates of scalp swelling/edema in all embryos on either diet (62.86% embryos exposed to NW8 diet, 22/35 embryos; 79.41% affected embryos exposed to NW16 diet, 27/34 embryos), which were suspected to be secondary to the embryo fixation, processing, and iodine contrasting, we also excluded these phenotypes from our anomaly counts. Although we noted protruding tongues in some embryos, these were not counted as anomalies. **Supplemental Data 1** has a comprehensive overview of anomalies in the embryos and **Supplemental Table 4** has an overview of regions assessed for anomalies.

### Analysis of kynurenine, NAD, and NADH by LC-HRMS

To determine the relative abundance of kynurenine in mouse serum, extracts were prepared and analyzed by high-resolution mass spectrometry (HRMS). Metabolites were extracted using cold 80/20 volume/volume (v/v) methanol/water. Samples were centrifuged at 17,000 g for 5 minutes at 4°C, and supernatants were transferred to clean tubes, followed by evaporation to dryness under nitrogen. Samples were reconstituted in 90/10 acetonitrile/water containing 1% formic acid, then 10 μL was injected into a Thermo Vanquish liquid chromatography (LC) system containing an Imtakt Intrada Amino Acid 2.1 x 150 mm column with 3 µm particle size. Mobile phase A (MPA) was acetonitrile containing 0.1% formic acid. Mobile phase B (MPB) was 50 mM ammonium formate. The flow rate was 250 µL/minute (at 35°C), and the gradient conditions were: initial 15% MPB, increased to 95% MPB at 20 minutes, held at 95% MPB for 5 minutes, returned to initial conditions and equilibrated for 5 minutes. The total run time was 30 minutes. Data were acquired using a Thermo Orbitrap Lumos Tribrid mass spectrometer under ESI positive ionization mode at a resolution of 240,000. The raw files were imported to Thermo Trace Finder software for final analysis.

To determine the relative abundance of NAD metabolites, extracts were prepared and analyzed by ultra-HRMS^70^. Approximately 60-80 mg of embryos were snap frozen in liquid nitrogen, then homogenized with Precellys Tissue Homogenizer. Metabolites were extracted using 1.0 mL ice-cold 40/40/20 volume/volume/volume (v/v/v) acetonitrile/methanol/water with 100mM formic acid, vortexed for 10 seconds, and allowed to sit on ice for 3 minutes. Then, 87 μL of 15% ammonium bicarbonate in water weight:volume (w:v) was added to neutralize the samples. After that, the mixture was incubated on ice for 20 minutes. Extracts were centrifuged at 17,000 *g* for 10 minutes at 4°C, and supernatants were transferred to autosampler vials for liquid chromatography (LC)-MS analysis. LC mobile phase A (MPA) was 95/5 (v/v) water/acetonitrile containing 20mM ammonium acetate and 20mM ammonium hydroxide (pH∼9), and mobile phase B (MPB) was acetonitrile. Thermo Vanquish LC system included a Xbridge BEH Amide column (3.5 µm particle size, 100 x 4.6 mm) with column compartment kept at 35°C. The autosampler tray was chilled to 4°C. The mobile phase flow rate was 350 µL/minute, and the gradient elution program was: 0-1 minutes, 85% MPB; 1-16 minutes, 85-15% MPB; 16-20 minutes, hold at 15% MPB, 20.5-25 minutes, 85% MPB. The total run time was 25 minutes. Data were acquired using a Thermo Orbitrap Exploris 240 Mass Spectrometer under ESI positive ionization mode at a resolution of 240,000. Raw data files were imported to Thermo Trace Finder software for final analysis, and the relative abundance of each metabolite was normalized by tissue weight.

### Untargeted metabolomics in E12.5 murine embryos

Untargeted metabolomics were performed on whole embryo homogenates using Thermo Scientific Vanquish UHPLC system (Thermo Fisher Scientific), which was equipped with a Vanquish Horizon Binary Pump H, a Vanquish Column Compartment H, and a Vanquish Split sampler HT. All the metabolites were separated using Hydrophilic Interaction Liquid Chromatography (HILIC) with an ACQUITY UPLC BEH Amide Column (130 Å, 1.7 µm, 2.1 mm X 150 mm) from Waters Corporation, as well as Reverse Phase (RP) chromatography utilizing an ACQUITY UPLC HSS T3 Column (100 Å, 1.8 µm, 2.1 mm X 150 mm), both in electro spray ionization (ESI) positive and negative modes as described in a previous publication^71^. The peak data obtained from both methods were normalized using median interquartile range (IQR). The differentially expressed metabolites were identified by two-way ANOVA with main effects for genotype (*Kmo^+/-^*and *Kmo^-/-^*) and diet (NW8 and NW16) with Tukey’s post hoc tests for multiple comparisons. Correction for false discovery was performed using the Benjamini-Hochberg method with a false discovery rate (FDR) threshold of 0.25 due to the exploratory nature of the analysis. Statistical testing was performed using R. For visualization, heat maps were generated using ggplot2^72^ and ComplexHeatmap^73^ packages in R. Metabolites were identified as having a significant genotype × diet interaction in the two-way ANOVA if they had an FDR ≤0.25 (**Supplemental Data 2**). The same metabolite(s) may be present in multiple heatmaps because they were significant for multiple factors in the two-way ANOVA. Pathway analysis was performed using differently expressed metabolites in all four embryo groups using MetaboAnalyst 6.0^50^.

### Statistical analyses

Male and female adult mice and embryos were used for phenotyping. Unpaired t-tests were used for all normally distributed two sample comparisons with equal variances or a Welch’s unpaired t-test if the variances were not equal. If data from two independent groups were nonparametric, a Mann-Whitney test was performed. A Fisher’s exact tests on the number of embryos with anomalies were used to determine the difference in the proportions of the groups. A p-value of ≤0.05 was considered statistically significant. All statistical analyses (except metabolomics) were performed using GraphPad Prism (version 11.0.0 (509)). Statistical analyses for the metabolomics data are provided in the previous sections.

### Study approval

The clinical studies described in this report were approved by the Institutional Review Boards (IRBs) at the National Institutes of Health and Baylor College of Medicine (BCM). The patient, one unaffected sister, and both parents were enrolled in the BCM site of the Undiagnosed Diseases Network (UDN). The youngest unaffected sister underwent clinical testing prenatally for the *KMO* variants and was not enrolled for any research evaluations. Informed consent was obtained prior to the initiation of any research procedures. Parents provided permission for the inclusion of clinical information about the deceased sister of the proband and for the reanalysis of her clinical exome data.

## Data Availability

All data produced in the present study are available upon reasonable request to the authors.

## Data availability

Raw and processed metabolomics data from murine samples are available from the authors upon request.

## Author contributions

N.M.A.-E. performed mouse experiments. D.G.L. and J.D.H. designed and generated the mouse model. S.L., J.A.R., E.L., S.E., C.G., and L.B. contributed clinical data. C.A.B. and B.L. supervised the human studies performed within the Undiagnosed Diseases Network. H.C. and S.D. performed and analyzed clinical targeted metabolomics data and provided feedback on mouse studies. P.L. contributed RNA sequencing data. N.M.A.-E., N.L.-V., C.H., R.M., A.E.C., T.L.R., and M.E.D. contributed to mouse phenotyping. N.M.A.-E., X.L., A.G., D.G.L., A.G., and H.Z. contributed to mouse genotyping. L.T., B.T., and P.L.L. contributed to mouse kynurenine and NAD data and analysis. P.N. and A.M.H.K. performed the murine untargeted metabolomics experiments. N.M.A.-E., A.K., C.C., V.P., and A.M. analyzed the untargeted metabolomics data and generated visuals. L.C.B. participated in and supervised mouse experiments and supervised clinical data collection and analysis. J.D.H. supervised mouse model designs and production. N.M.A.-E. prepared the initial draft of the manuscript with L.C.B. and C.H.. All authors reviewed and provided edits to the final version of the manuscript.

## Acknowledgements

We thank the patient and family for participating in our research studies. We acknowledge the Medical Genetics Multiomics Laboratory (MGML) for generating the RNA-seq data. This work is the result of NIH funding, in part, and is subject to the NIH Public Access Policy. Through acceptance of this federal funding, the NIH has been given a right to make the work publicly available in PubMed Central. We gratefully acknowledge the Victor Chang Cardiac Research Institute Innovation Centre, funded by the NSW Government, as well as funding from the Freedman Foundation for the Metabolomics Facility.

## Web Resources

CADD, https://cadd.gs.washington.edu/

Clinvar, https://www.ncbi.nlm.nih.gov/clinvar/

gnomAD, https://gnomad.broadinstitute.org/

GraphPad Prism, www.graphpad.com/

OMIM, https://www.omim.org/

Mouse Genome Informatics (MGI), https://www.informatics.jax.org/

REVEL, https://sites.google.com/site/revelgenomics/

Wellcome Sanger Institute Genome Editing (WGE), https://wge.stemcell.sanger.ac.uk/

Synthego Inference of CRISPR Edits (ICE), https://ice.synthego.com/#/

SnapGene® software, from Dotmatics; available at snapgene.com

Slicer 3D, https://www.slicer.org/

MetaboAnalyst, https://www.metaboanalyst.ca/home.xhtml

## Supplemental Figures

**Supplemental Figure 1. RNA sequencing of a whole blood sample from II-2 reveals altered splicing of *KMO.*** Exome data demonstrates the location of the NM_003679.4:c.223-1G>T variant in red and highlighted in yellow. RNA sequencing demonstrates loss of the canonical splice acceptor site in exon 4 (highlighted in yellow) with gain of a new acceptor site (dashed line and arrow) leading to loss of the first 13 nucleotides of exon 4 in a subset of reads. The sashimi plot demonstrates the four reads (red box) supporting the cryptic splice junction.

**Supplemental Figure 2. Design and validation of *Kmo* deletion.** A) CRISPR strategy for the *Kmo* knockout mouse model, including guide sites, primers for genotyping and RT-qPCR (see **Supplemental Table 3** for sequences), and predicted frameshift caused by splicing from exon 4 to 6 and early stop in downstream exon. White boxes with outline = untranslated sequence; blue filled boxes = coding sequence; yellow boxes = frameshifted sequence; * = premature stop. B) Sanger sequencing of edited *Kmo* breakpoint demonstrating expected deletion. Sequences aligned using GRCm39 and NM_133809.1.

**Supplemental Figure 3. DORV detected in an E18.5 *Kmo^-/-^*embryo exposed to the NW8 diet.** A) Representative images from coronal plane demonstrating a morphologically normal heart in a *Kmo^+/-^* embryo (ID=6.3). B) Images from coronal plane demonstrating DORV in a *Kmo^-/-^*embryo (ID=2.5). C) Bar graphs of percentages of embryos with at least one anomaly in the given region/organ per genotype-diet group. A-B) Embryo IDs listed in parentheses correspond to embryos that can be found in **Supplemental Data 1,** Sheet 1.

**Supplemental Figure 4. Crown-rump lengths of E18.5 embryos and E12.5 weights.** A-B) Only *Kmo^-/-^* embryos derived from *Kmo^-/-^*dams fed the NW8 diet have significantly shorter crown-rump lengths than their heterozygous littermates. Mann-Whitney tests, error bars represent mean and SD. C-D) E12.5 embryo weights did not differ significantly between genotypes or diet treatments. Unpaired t-tests, error bars represent mean and SD.

**Supplemental Figure 5. µCT bone measurements of E18.5 embryos.** A-B) Only *Kmo^-/-^* embryos derived from *Kmo^-/-^*dams fed the NW8 diet have significantly shorter ulnas than their heterozygous littermates. C-D) *Kmo^-/-^* embryos derived from *Kmo^-/-^*dams fed the NW8 diet have trends for shorter femurs, but *Kmo^-/-^*embryos and *Kmo^+/-^* embryos derived from *Kmo^-/-^*dams fed the NW16 diet do not demonstrate a trend for shorter femurs. E-F) *Kmo^-/-^* embryos derived from *Kmo^-/-^* dams fed the NW8 diet demonstrate a trend for shorter radii as compared to their heterozygous littermates. *Kmo^-/-^* embryos and *Kmo^+/-^* embryos derived from *Kmo^-/-^* dams fed the NW16 diet do not demonstrate a trend for shorter radii. A-D) Welch’s t-tests, n = 13 *Kmo^+/-^*and n = 14 *Kmo^-/-^* embryos exposed to NW8 diet; n = 17 *Kmo^+/-^*and n = 17 *Kmo^-/-^* embryos exposed to NW16 diet. Error bars represent mean and SD.

**Supplemental Figure 6. Heatmap of metabolites that were significant for interaction (genotype × diet) effects detected in mouse embryos through untargeted metabolomics.** Data demonstrate that *Kmo^-/-^* embryos exposed to the NW8 diet *in utero* have different patterns of relative metabolite abundance as compared to embryos on a control diet or heterozygous littermates. The same metabolite(s) may be present in multiple heatmaps because they were significant for multiple factors in the two-way ANOVA. Data shown is also presented as dot plots for individual metabolites in Supplemental Figure 11 and extended Supplemental Figure 11.

**Supplemental Figure 7. Heatmap of metabolites that were significant for diet effects detected in mouse embryos through untargeted metabolomics.** Data demonstrate that embryos exposed to a low niacin diet *in utero* have different patterns of relative metabolite abundance as compared to embryos on a control diet. The same metabolite(s) may be present in multiple heatmaps because they were significant for multiple factors in the two-way ANOVA.

**Supplemental Figure 8. Heatmap of metabolites that were significant for genotype effects detected in mouse embryos through untargeted metabolomics.** Data demonstrate that few metabolites clustered differently depending on embryo genotype. The same metabolite(s) may be present in multiple heatmaps because they were significant for multiple factors in the two-way ANOVA.

**Supplemental Figure 9 and extended Supplemental Figure 9. Dot plots of metabolites that were significant for interaction (genotype × diet) effects detected in mouse embryos through untargeted metabolomics.** Some metabolite data (NAD^+^, cADPR, dGDP, and thymidine) are also shown in **Figure 6**. Data shown is also present as a heatmap in Supplemental Figure 8 and the two-way ANOVA and Tukey’s post hoc results are available in **Supplemental Data 2**. Error bars represent maximum to minimum values, black circles represent mean values.

## Supplemental Tables

**Supplemental Table 1.** Levels of metabolites of the NAD metabolome are altered in whole blood from the proband (II-2). Values from NAD^+^, NMN, and excretion products were excluded from the analysis because the proband was taking vitamin B3 supplements during the sample collection.

**Supplemental Table 2.** Adult *Kmo^+/-^* and *Kmo^-/-^* mice were observed at the expected Mendelian ratio. E18.5 and E12.5 *Kmo^+/-^* and *Kmo^-/-^* embryos on the NW8 and NW16 diets were observed at the expected Mendelian ratio. The sex distributions for E18.5 embryos on the NW8 and NW16 diets were also observed at the expected ratio.

**Supplemental Table 3.** Genotyping, *Sry*, and RT-qPCR primers.

**Supplemental Table 4.** Anatomical areas in murine embryos assessed for congenital anomalies by µCT.

## Supplemental Data (Excel files)

**Supplemental Data 1.** Sheet 1: Comprehensive heatmap of anatomical areas screened in murine embryos and anomalies detected per embryo. Red boxes in the heatmap indicate that researchers noted a phenotype for the given category/tissue in an embryo. Scalp swelling or edema, hydronephrosis, and protruding tongues were noted but not counted as anomalies. Please see Methods and Materials for further details on phenotyping. Data from this sheet was used to generate panels in **Figure 4**. Sheet 2: Tables using phenotypes from Sheet 1 to list murine embryo anomalies detected per general anatomic regions. Data from Sheet 2 was used to generate **Supplemental Figure 5C**.

**Supplemental Data 2.** Results from two-way ANOVA and Tukey’s post-hoc test for multiple comparisons for E12.5 embryo metabolomics. On sheet “2-Way ANOVA,” columns titled “Diet_p”, “Genotype_p”, and “Interaction_p” contain raw p-values from two-way ANOVA. Columns titled “Diet_FDR”, “Genotype_FDR”, and “Interaction_FDR” are the Benjamini-Hochberg (BH) FDR adjusted p-values; BH FDR p-values ≤0.25 were considered significant.

## Bibliography

1. Xie, N., Zhang, L., Gao, W., Huang, C., Huber, P.E., Zhou, X., Li, C., Shen, G., and Zou, B. (2020). NAD+ metabolism: pathophysiologic mechanisms and therapeutic potential. Signal Transduction and Targeted Therapy.

2. Poyan Mehr, A., Tran, M.T., Ralto, K.M., Leaf, D.E., Washco, V., Messmer, J., Lerner, A., Kher, A., Kim, S.H., Khoury, C.C., et al. (2018). De novo NAD+ biosynthetic impairment in acute kidney injury in humans. Nature Medicine 24, 1351–1359. 10.1038/s41591-018-0138-z.

3. Ralto, K.M., Rhee, E.P., and Parikh, S.M. (2020). NAD+ homeostasis in renal health and disease. Nature Reviews Nephrology.

4. Warren, A., Porter, R.M., Reyes-Castro, O., Ali, M.M., Marques-Carvalho, A., Kim, H.-N., Gatrell, L.B., Schipani, E., Nookaew, I., O’Brien, C.A., et al. (2023). The NAD salvage pathway in mesenchymal cells is indispensable for skeletal development in mice. Nature Communications 14, 3616–3616. 10.1038/s41467-023-39392-7.

5. Castro-Portuguez, R., and Sutphin, G.L. (2020). Kynurenine pathway, NAD+ synthesis, and mitochondrial function: Targeting tryptophan metabolism to promote longevity and healthspan. Experimental Gerontology. 2020/01/16 ed.

6. Fukuwatari, T., and Shibata, K. (2013). Nutritional aspect of tryptophan metabolism. International Journal of Tryptophan Research.

7. Liu, L., Su, X., Quinn, W.J., Hui, S., Krukenberg, K., Frederick, D.W., Redpath, P., Zhan, L., Chellappa, K., White, E., et al. (2018). Quantitative Analysis of NAD Synthesis-Breakdown Fluxes. Cell Metabolism 27, 1067–1080.e1065. 10.1016/j.cmet.2018.03.018.

8. Zapata-Pérez, R., Wanders, R.J.A., Karnebeek, C.D.M., and Houtkooper, R.H. (2021). NAD + homeostasis in human health and disease. EMBO Molecular Medicine 13, e13943–e13943. 10.15252/emmm.202113943.

9. Katsyuba, E., Romani, M., Hofer, D., and Auwerx, J. (2020). NAD+ homeostasis in health and disease. Nature Metabolism 2, 9–31. 10.1038/s42255-019-0161-5.

10. Shi, H., Enriquez, A., Rapadas, M., Martin, E.M.M.A., Wang, R., Moreau, J., Lim, C.K., Szot, J.O., Ip, E., Hughes, J.N., et al. (2017). NAD Deficiency, Congenital Malformations, and Niacin Supplementation. New England Journal of Medicine 377, 544–552. 10.1056/nejmoa1616361.

11. Szot, J.O., Campagnolo, C., Cao, Y., Iyer, K.R., Cuny, H., Drysdale, T., Flores-Daboub, J.A., Bi, W., Westerfield, L., Liu, P., et al. (2020). Bi-allelic Mutations in NADSYN1 Cause Multiple Organ Defects and Expand the Genotypic Spectrum of Congenital NAD Deficiency Disorders. American Journal of Human Genetics 106, 129–136. 10.1016/j.ajhg.2019.12.006.

12. Szot, J.O., Cuny, H., Martin, E.M.M.A., Sheng, D.Z., Iyer, K., Portelli, S., Nguyen, V., Gereis, J.M., Alankarage, D., Chitayat, D., et al. (2024). A metabolic signature for NADSYN1-dependent congenital NAD deficiency disorder. The Journal of Clinical Investigation 134. 10.1172/JCI174824.

13. Szot, J.O., Slavotinek, A., Chong, K., Brandau, O., Nezarati, M., Cueto-González, A.M., Patel, M.S., Devine, W.P., Rego, S., Acyinena, A.P., et al. (2021). New cases that expand the genotypic and phenotypic spectrum of Congenital NAD Deficiency Disorder. Human Mutation 42, 862–876. 10.1002/humu.24211.

14. Erbs, E., Brasen, C.L., Lund, A.M., and Rasmussen, M. (2023). Adult patient diagnosed with NADSYN1 associated congenital NAD deficiency and analysis of NAD levels to be published in: European Journal of Medical Genetics. European Journal of Medical Genetics 66, 104698–104698. 10.1016/j.ejmg.2023.104698.

15. Ehmke, N., Cusmano-Ozog, K., Koenig, R., Holtgrewe, M., Nur, B., Mihci, E., Babcock, H., Gonzaga-Jauregui, C., Overton, J.D., Xiao, J., et al. (2020). Biallelic variants in KYNU cause a multisystemic syndrome with hand hyperphalangism. Bone 133, 115219–115219. 10.1016/j.bone.2019.115219.

16. Kortbawi, H., Ames, E., Pritchard, A., Devine, P., van Ziffle, J., and Slavotinek, A. (2022). Further description of two patients with biallelic variants in NADSYN1 in association with cardiac and vertebral anomalies. American Journal of Medical Genetics, Part A 188, 2479–2484. 10.1002/ajmg.a.62765.

17. Lin, J., Zhao, L., Zhao, S., Li, S., Zhao, Z., Chen, Z., Zheng, Z., Shao, J., Niu, Y., Li, X., et al. (2021). Disruptive NADSYN1 Variants Implicated in Congenital Vertebral Malformations. Genes 12, 1615.

18. Schüle, I., Berger, U., Matysiak, U., Ruzaike, G., Stiller, B., Pohl, M., Spiekerkoetter, U., Lausch, E., Grünert, S.C., and Schmidts, M. (2021). A Homozygous Deletion of Exon 5 of KYNU Resulting from a Maternal Chromosome 2 Isodisomy (UPD2) Causes Catel-Manzke-Syndrome/VCRL Syndrome. Genes 12, 879.

19. Mark, P., and Dunwoodie, S. (2023). Congenital NAD Deficiency Disorder. Synonym: Vertebral, Cardiac, Renal, and Limb Defects (VCRL). In F.J. Adam MP, Mirzaa GM, et al., editors. ed. GeneReviews® [Internet].

20. Aubert-Mucca, M., Janel, C., Porquet-Bordes, V., Patat, O., Touraine, R., Edouard, T., Michot, C., Tessier, A., Cormier-Daire, V., Attie-Bitach, T., and Baujat, G. (2023). Clinical heterogeneity of NADSYN1-associated VCRL syndrome. Clinical Genetics 104, 114–120. 10.1111/cge.14328.

21. Ahmed, Z., Thahiem, S., Bakhsh, A., Khan, M.J., Umair, M., and Khan, H. (2026). Homozygous Variant in NADSYN1 Causes Multiple Congenital Vertebral Malformation, With Neurodevelopmental Disorder. International Journal of Developmental Neuroscience 86, e70140. 10.1002/jdn.70140.

22. Mark, P.R. (2022). NAD+ deficiency in human congenital malformations and miscarriage: A new model of pleiotropy. American Journal of Medical Genetics, Part A 188, 2834–2849. 10.1002/ajmg.a.62764.

23. Bozon, K., Cuny, H., Sheng, D.Z., Martin, E.M.M.A., Sipka, A., Young, P., Humphreys, D.T., and Dunwoodie, S.L. (2025). Impaired yolk sac NAD metabolism disrupts murine embryogenesis with relevance to human birth defects. eLife 13, RP97649. 10.7554/eLife.97649.

24. Cuny, H., Rapadas, M., Gereis, J., Martin, E.M.M.A., Kirk, R.B., Shi, H., and Dunwoodie, S.L. (2020). NAD deficiency due to environmental factors or gene–environment interactions causes congenital malformations and miscarriage in mice. Proceedings of the National Academy of Sciences of the United States of America 117, 3738–3747. 10.1073/pnas.1916588117.

25. Dunwoodie, S.L., Bozon, K., Szot, J.O., and Cuny, H. (2023). Nicotinamide Adenine Dinucleotide Deficiency and Its Impact on Mammalian Development. Antioxidants & Redox Signaling 39, 1108–1132. 10.1089/ars.2023.0349.

26. Amaral, M., Levy, C., Heyes, D.J., Lafite, P., Outeiro, T.F., Giorgini, F., Leys, D., and Scrutton, N.S. (2013). Structural basis of kynurenine 3-monooxygenase inhibition. Nature 496, 382–385. 10.1038/nature12039.

27. Hughes, T.D., Güner, O.F., Iradukunda, E.C., Phillips, R.S., and Bowen, J.P. (2022). The Kynurenine Pathway and Kynurenine 3-Monooxygenase Inhibitors. Molecules 27. 10.3390/molecules27010273.

28. Maddison, D.C., Alfonso-Núñez, M., Swaih, A.M., Breda, C., Campesan, S., Allcock, N., Straatman-Iwanowska, A., Kyriacou, C.P., and Giorgini, F. (2020). A novel role for kynurenine 3-monooxygenase in mitochondrial dynamics. PLoS Genetics 16, e1009129–e1009129. 10.1371/JOURNAL.PGEN.1009129.

29. Guez, J., Goodrich, J.K., Moldovan, M.A., Chao, K.R., Kar, P., Panchal, R., Wilson, M.W., Laricchia, K.M., Rohlicek, G., Biba, D., et al. (2026). Integrating 730,947 exome sequences with clinical literature improves gene discovery. medRxiv, 2026.2003.2023.26349081. 10.64898/2026.03.23.26349081.

30. Jaganathan, K., Kyriazopoulou Panagiotopoulou, S., McRae, J.F., Darbandi, S.F., Knowles, D., Li, Y.I., Kosmicki, J.A., Arbelaez, J., Cui, W., Schwartz, G.B., et al. (2019). Predicting Splicing from Primary Sequence with Deep Learning. Cell 176, 535–548.e524. 10.1016/j.cell.2018.12.015.

31. de Sainte Agathe, J.-M., Filser, M., Isidor, B., Besnard, T., Gueguen, P., Perrin, A., Van Goethem, C., Verebi, C., Masingue, M., Rendu, J., et al. (2023). SpliceAI-visual: a free online tool to improve SpliceAI splicing variant interpretation. Human Genomics 17, 7. 10.1186/s40246-023-00451-1.

32. Rentzsch, P., Witten, D., Cooper, G.M., Shendure, J., and Kircher, M. (2019). CADD: predicting the deleteriousness of variants throughout the human genome. Nucleic Acids Research 47, D886–D894. 10.1093/nar/gky1016.

33. Rentzsch, P., Schubach, M., Shendure, J., and Kircher, M. (2021). CADD-Splice—improving genome-wide variant effect prediction using deep learning-derived splice scores. Genome Medicine 13, 31. 10.1186/s13073-021-00835-9.

34. Sim, N.-L., Kumar, P., Hu, J., Henikoff, S., Schneider, G., and Ng, P.C. (2012). SIFT web server: predicting effects of amino acid substitutions on proteins. Nucleic Acids Research 40, W452–W457. 10.1093/nar/gks539.

35. Adzhubei, I.A., Schmidt, S., Peshkin, L., Ramensky, V.E., Gerasimova, A., Bork, P., Kondrashov, A.S., and Sunyaev, S.R. (2010). A method and server for predicting damaging missense mutations. Nature Methods 7, 248–249. 10.1038/nmeth0410-248.

36. Schwarz, J.M., Cooper, D.N., Schuelke, M., and Seelow, D. (2014). MutationTaster2: mutation prediction for the deep-sequencing age. Nature Methods 11, 361–362. 10.1038/nmeth.2890.

37. Tordai, H., Torres, O., Csepi, M., Padányi, R., Lukács, G.L., and Hegedűs, T. (2024). Analysis of AlphaMissense data in different protein groups and structural context. Scientific Data 11, 495. 10.1038/s41597-024-03327-8.

38. Cheng, J., Novati, G., Pan, J., Bycroft, C., Žemgulytė, A., Applebaum, T., Pritzel, A., Wong, L.H., Zielinski, M., Sargeant, T., et al. Accurate proteome-wide missense variant effect prediction with AlphaMissense. Science 381, eadg7492. 10.1126/science.adg7492.

39. Ioannidis, N.M., Rothstein, J.H., Pejaver, V., Middha, S., McDonnell, S.K., Baheti, S., Musolf, A., Li, Q., Holzinger, E., Karyadi, D., et al. (2016). REVEL: An Ensemble Method for Predicting the Pathogenicity of Rare Missense Variants. The American Journal of Human Genetics 99, 877–885. 10.1016/j.ajhg.2016.08.016.

40. Kim, H.T., Na, B.K., Chung, J., Kim, S., Kwon, S.K., Cha, H., Son, J., Cho, J.M., and Hwang, K.Y. (2018). Structural Basis for Inhibitor-Induced Hydrogen Peroxide Production by Kynurenine 3-Monooxygenase. Cell Chemical Biology 25, 426–438.e424. 10.1016/j.chembiol.2018.01.008.

41. Hutchinson, J.P., Rowland, P., Taylor, M.R.D., Christodoulou, E.M., Haslam, C., Hobbs, C.I., Holmes, D.S., Homes, P., Liddle, J., Mole, D.J., et al. (2017). Structural and mechanistic basis of differentiated inhibitors of the acute pancreatitis target kynurenine-3-monooxygenase. Nature Communications 8, 15827. 10.1038/ncomms15827.

42. Mimasu, S., Yamagishi, H., Kubo, S., Kiyohara, M., Matsuda, T., Yahata, T., Thomson, H.A., Hupp, C.D., Liu, J., Okuda, T., and Kakefuda, K. (2021). Full-length in meso structure and mechanism of rat kynurenine 3-monooxygenase inhibition. Communications Biology 4, 159. 10.1038/s42003-021-01666-5.

43. Rossi, F., Miggiano, R., Ferraris, D.M., and Rizzi, M. (2019). The Synthesis of Kynurenic Acid in Mammals: An Updated Kynurenine Aminotransferase Structural KATalogue. Frontiers in Molecular Biosciences Volume 6 - 2019.

44. Lonsdale, J., Thomas, J., Salvatore, M., Phillips, R., Lo, E., Shad, S., Hasz, R., Walters, G., Garcia, F., Young, N., et al. (2013). The Genotype-Tissue Expression (GTEx) project. Nature Genetics 45, 580–585. 10.1038/ng.2653.

45. Giorgini, F., Huang, S.Y., Sathyasaikumar, K.V., Notarangelo, F.M., Thomas, M.A.R., Tararina, M., Wu, H.Q., Schwarcz, R., and Muchowski, P.J. (2013). Targeted deletion of kynurenine 3-Monooxygenase in mice a new tool for studying kynurenine pathway metabolism in periphery and brain. Journal of Biological Chemistry 288, 36554–36566. 10.1074/jbc.M113.503813.

46. Dumont, K.D., Jannig, P.R., Porsmyr-Palmertz, M., and Ruas, J.L. (2025). Constitutive loss of kynurenine-3-monooxygenase changes circulating kynurenine metabolites without affecting systemic energy metabolism. American Journal of Physiology-Endocrinology and Metabolism 328, E274–E285. 10.1152/ajpendo.00386.2024.

47. Erhardt, S., Pocivavsek, A., Repici, M., Liu, X.C., Imbeault, S., Maddison, D.C., Thomas, M.A.R., Smalley, J.L., Larsson, M.K., Muchowski, P.J., et al. (2017). Adaptive and Behavioral Changes in Kynurenine 3-Monooxygenase Knockout Mice: Relevance to Psychotic Disorders. Biological Psychiatry 82, 756–765. 10.1016/j.biopsych.2016.12.011.

48. Elmore, S.A., Kavari, S.L., Hoenerhoff, M.J., Mahler, B., Scott, B.E., Yabe, K., and Seely, J.C. (2019). Histology Atlas of the Developing Mouse Urinary System With Emphasis on Prenatal Days E10.5-E18.5. Toxicologic Pathology 47, 865–886. 10.1177/0192623319873871.

49. Andrés-Delgado, L., and Mercader, N. (2016). Interplay between cardiac function and heart development. Biochimica et Biophysica Acta (BBA) - Molecular Cell Research 1863, 1707–1716. 10.1016/j.bbamcr.2016.03.004.

50. Pang, Z., Lu, Y., Zhou, G., Hui, F., Xu, L., Viau, C., Spigelman, Aliya F., MacDonald, Patrick E., Wishart, David S., Li, S., and Xia, J. (2024). MetaboAnalyst 6.0: towards a unified platform for metabolomics data processing, analysis and interpretation. Nucleic Acids Research 52, W398–W406. 10.1093/nar/gkae253.

51. Cuny, H., Shand, A.W., Goth, J., Sheng, D.Z., Tossey, T., Martin, E.M.M.A., Sipka, A., Aleshin, O., Schneuer, F.J., Nassar, N., and Dunwoodie, S.L. (2025). Identification of potential NAD-related biomarkers of recurrent miscarriage risk. Human Reproduction 40, 2247–2259. 10.1093/humrep/deaf195.

52. Bozon, K., Cuny, H., Sunn, N., Martin, E.M.M.A., Sheng, D.Z., Chapman, G., and Dunwoodie, S.L. (2026). Timing of NAD Deficiency During Organogenesis Dictates Defect Type and Penetrance. The FASEB Journal 40, e71504. 10.1096/fj.202502824RRR.

53. Palzer, L., Bader, J.J., Angel, F., Witzel, M., Blaser, S., McNeil, A., Wandersee, M.K., Leu, N.A., Lengner, C.J., Cho, C.E., et al. (2018). Alpha-Amino-Beta-Carboxy-Muconate-Semialdehyde Decarboxylase Controls Dietary Niacin Requirements for NAD+ Synthesis. Cell Reports 25, 1359–1370.e1354. 10.1016/j.celrep.2018.09.091.

54. Ikeda, M., Tsuji, H., Nakamura, S., Ichiyama, A., Nishizuka, Y., and Hayaishi, O. (1965). STUDIES ON THE BIOSYNTHESIS OF NICOTINAMIDE ADENINE DINUCLEOTIDE. II. A ROLE OF PICOLINIC CARBOXYLASE IN THE BIOSYNTHESIS OF NICOTINAMIDE ADENINE DINUCLEOTIDE FROM TRYPTOPHAN IN MAMMALS. The Journal of biological chemistry 240, 1395–1401.

55. Mori, Y., Mouri, A., Kunisawa, K., Hirakawa, M., Kubota, H., Kosuge, A., Niijima, M., Hasegawa, M., Kurahashi, H., Murakami, R., et al. (2021). Kynurenine 3-monooxygenase deficiency induces depression-like behavior via enhanced antagonism of α7 nicotinic acetylcholine receptors by kynurenic acid. Behavioural Brain Research 405, 113191–113191. 10.1016/j.bbr.2021.113191.

56. Koscielny, G., Yaikhom, G., Iyer, V., Meehan, T.F., Morgan, H., Atienza-Herrero, J., Blake, A., Chen, C.-K., Easty, R., Di Fenza, A., et al. (2014). The International Mouse Phenotyping Consortium Web Portal, a unified point of access for knockout mice and related phenotyping data. Nucleic Acids Research 42, D802–D809. 10.1093/nar/gkt977.

57. Tashiro, T., Murakami, Y., Mouri, A., Imamura, Y., Nabeshima, T., Yamamoto, Y., and Saito, K. (2017). Kynurenine 3-monooxygenase is implicated in antidepressants-responsive depressive-like behaviors and monoaminergic dysfunctions. Behavioural Brain Research 317, 279–285. 10.1016/j.bbr.2016.09.050.

58. Goorden, S.M.I., van Haaften-Visser, D.Y., Trętowicz, M.M., Bonte, R., Bogaerts, E., Jamal, Y., Vrieswijk, S., Huijser, E., Bökenkamp, R., van der Palen, R.L.F., et al. (2025). Two new cases of KYNU deficiency: Further delineation of the phenotypic and biochemical spectrum and exploration of treatment options. Molecular Genetics and Metabolism 146, 109194. 10.1016/j.ymgme.2025.109194.

59. Yang, Y., Muzny, D.M., Xia, F., Niu, Z., Person, R., Ding, Y., Ward, P., Braxton, A., Wang, M., Buhay, C., et al. (2014). Molecular Findings Among Patients Referred for Clinical Whole-Exome Sequencing. JAMA 312, 1870–1879. 10.1001/jama.2014.14601.

60. Karczewski, K.J., Francioli, L.C., Tiao, G., Cummings, B.B., Alföldi, J., Wang, Q., Collins, R.L., Laricchia, K.M., Ganna, A., Birnbaum, D.P., et al. (2020). The mutational constraint spectrum quantified from variation in 141,456 humans. Nature 581, 434–443. 10.1038/s41586-020-2308-7.

61. Murdock, D.R., Dai, H., Burrage, L.C., Rosenfeld, J.A., Ketkar, S., Müller, M.F., Yépez, V.A., Gagneur, J., Liu, P., Chen, S., et al. (2020). Transcriptome-directed analysis for Mendelian disease diagnosis overcomes limitations of conventional genomic testing. The Journal of Clinical Investigation 131. 10.1172/JCI141500.

62. Zhao, S., Macakova, K., Sinson, J.C., Dai, H., Rosenfeld, J., Zapata, G.E., Li, S., Ward, P.A., Wang, C., Qu, C., et al. (2025). Clinical validation of RNA sequencing for Mendelian disorder diagnostics. The American Journal of Human Genetics 112, 779–792. 10.1016/j.ajhg.2025.02.006.

63. Hodgkins, A., Farne, A., Perera, S., Grego, T., Parry-Smith, D.J., Skarnes, W.C., and Iyer, V. (2015). WGE: a CRISPR database for genome engineering. Bioinformatics 31, 3078–3080. 10.1093/bioinformatics/btv308.

64. Lanza, D.G., Mao, J., Lorenzo, I., Liao, L., Seavitt, J.R., Ljungberg, M.C., Simpson, E.M., DeMayo, F.J., and Heaney, J.D. (2024). An oocyte-specific Cas9-expressing mouse for germline CRISPR/Cas9-mediated genome editing. Genesis 62, e23589. 10.1002/dvg.23589.

65. Livak, K.J., and Schmittgen, T.D. (2001). Analysis of Relative Gene Expression Data Using Real-Time Quantitative PCR and the 2−ΔΔCT Method. Methods 25, 402–408. 10.1006/meth.2001.1262.

66. Hsu, C.-W., Kalaga, S., Akoma, U., Rasmussen, T.L., Christiansen, A.E., and Dickinson, M.E. (2019). High Resolution Imaging of Mouse Embryos and Neonates with X-Ray Micro-Computed Tomography. Current Protocols in Mouse Biology 9, e63–e63. 10.1002/cpmo.63.

67. Dickinson, M.E., Flenniken, A.M., Ji, X., Teboul, L., Wong, M.D., White, J.K., Meehan, T.F., Weninger, W.J., Westerberg, H., Adissu, H., et al. (2016). High-throughput discovery of novel developmental phenotypes. Nature 537, 508–514. 10.1038/nature19356.

68. Fedorov, A., Beichel, R., Kalpathy-Cramer, J., Finet, J., Fillion-Robin, J.-C., Pujol, S., Bauer, C., Jennings, D., Fennessy, F., Sonka, M., et al. (2012). 3D Slicer as an image computing platform for the Quantitative Imaging Network. Magnetic Resonance Imaging 30, 1323–1341. 10.1016/j.mri.2012.05.001.

69. Springer, D.A., Allen, M., Hoffman, V., Brinster, L., Starost, M.F., Bryant, M., and Eckhaus, M. (2014). Investigation and identification of etiologies involved in the development of acquired hydronephrosis in aged laboratory mice with the use of high-frequency ultrasound imaging. Pathobiology of Aging & Age-related Diseases 4, 24932. 10.3402/pba.v4.24932.

70. Lu, W., Wang, L., Chen, L., Hui, S., and Rabinowitz, J.D. (2018). Extraction and Quantitation of Nicotinamide Adenine Dinucleotide Redox Cofactors. Antioxidants & Redox Signaling 28, 167–179. 10.1089/ars.2017.7014.

71. Kamal, A.H.M., Putluri, V., Gandhi, T., Ambati, C.S.R., Amara, C.S., Kami Reddy, K.R., Spradlin, M.L., Koirala, A., Jorvekar, S.B., Grimm, S.L., et al. (2026). Leveraging untargeted metabolomics in combination with machine learning to uncover novel insights into bladder cancer. Cancer & Metabolism 14, 8. 10.1186/s40170-026-00427-4.

72. Wickham, H. (2016). ggplot2: Elegant Graphics for Data Analysis (Springer-Verlag New York). 10.1007/978-3-319-24277-4_9.

73. Gu, Z., Eils, R., and Schlesner, M. (2016). Complex heatmaps reveal patterns and correlations in multidimensional genomic data. Bioinformatics 32, 2847–2849. 10.1093/bioinformatics/btw313.

74. Goldsmith, G.A. (1958). Niacin-Tryptophan Relationships in Man and Niacin Requirement. The American Journal of Clinical Nutrition 6, 479–486. 10.1093/ajcn/6.5.479.

75. Bachmanov, A.A., Reed, D.R., Beauchamp, G.K., and Tordoff, M.G. (2002). Food Intake, Water Intake, and Drinking Spout Side Preference of 28 Mouse Strains. Behavior Genetics 32, 435–443. 10.1023/A:1020884312053.

