## Supplemental materials for "Biallelic Variants in *KMO* Cause a Novel Form of Congenital NAD Deficiency"

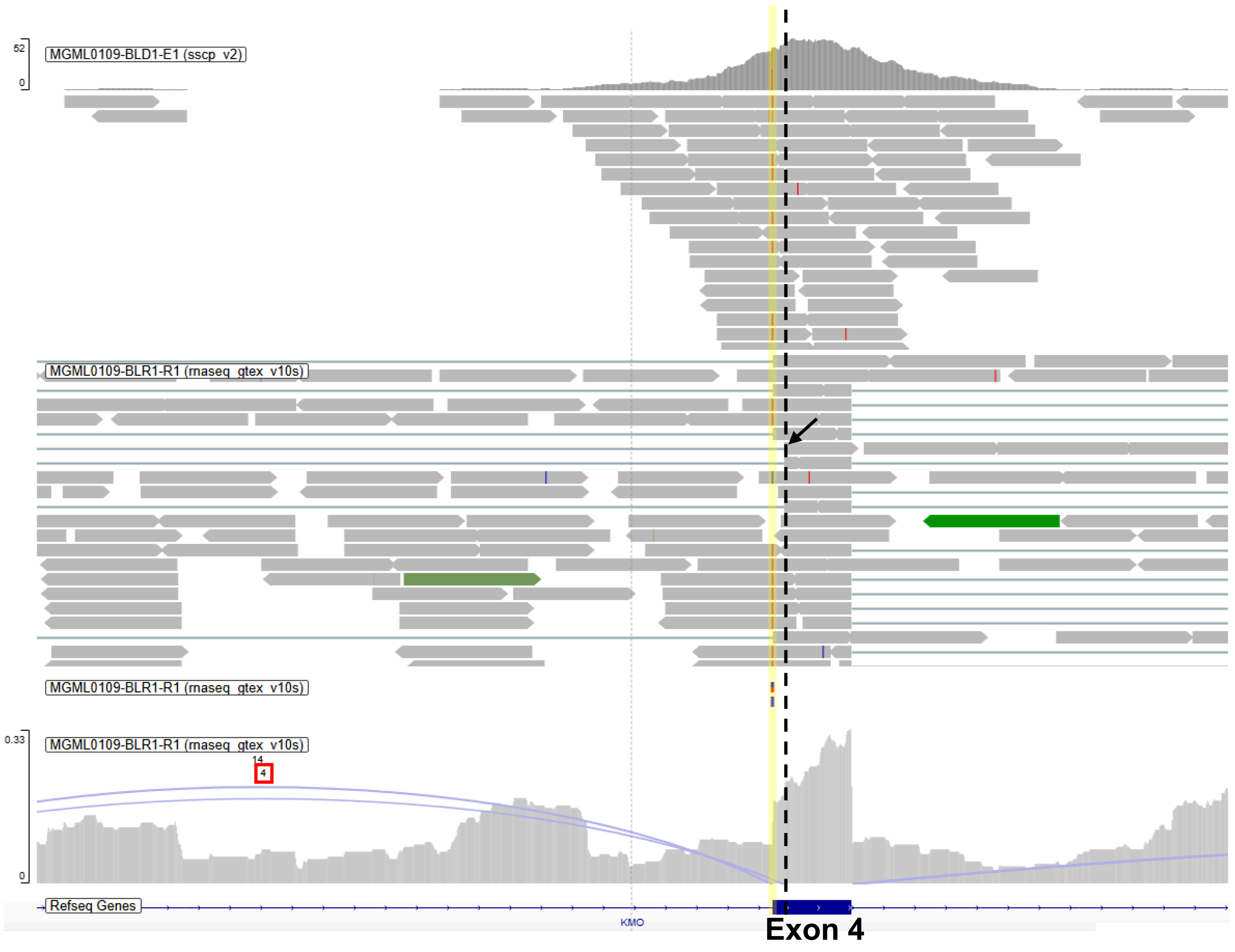


### Supplemental Figure 1

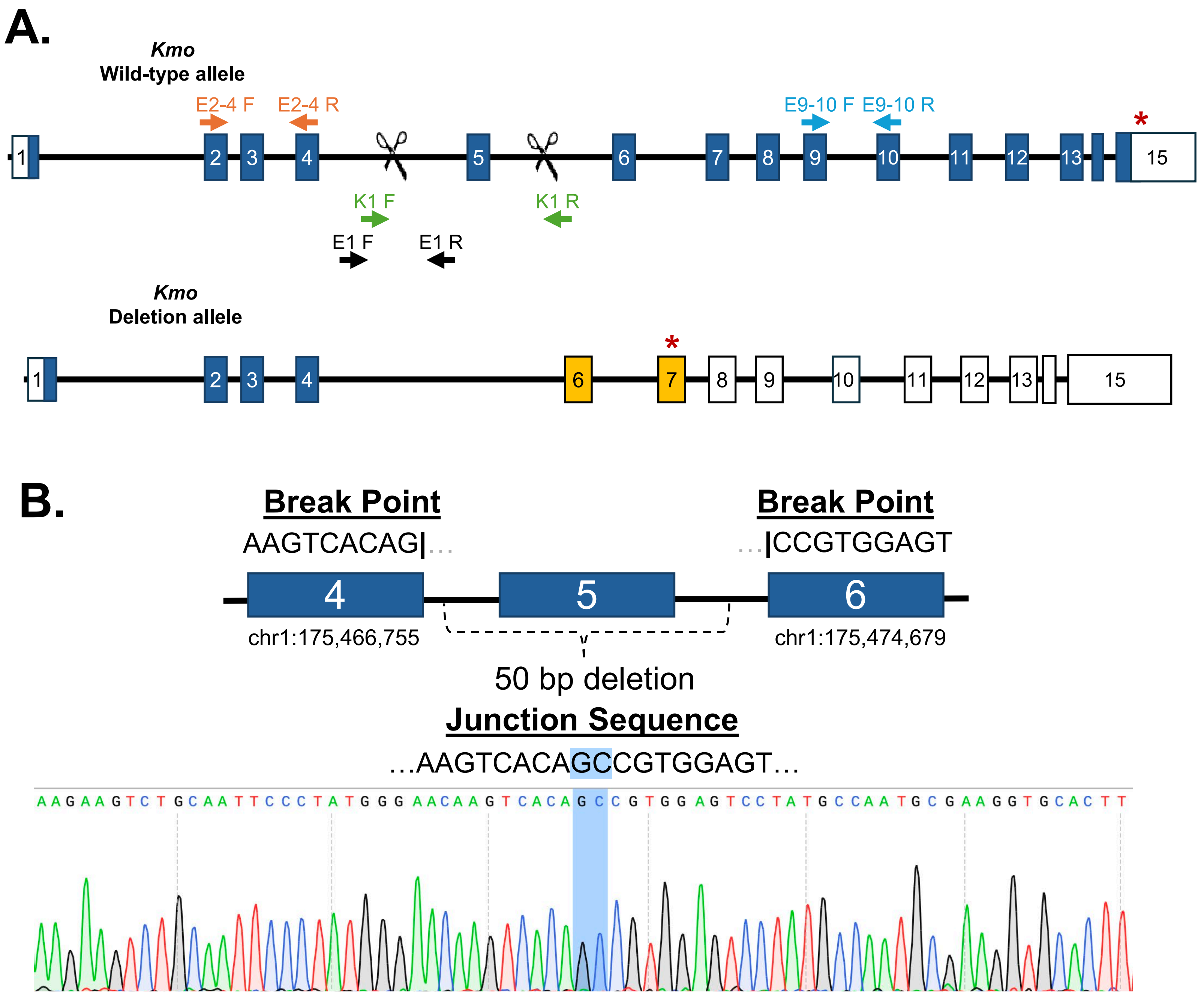


### Supplemental Figure 2

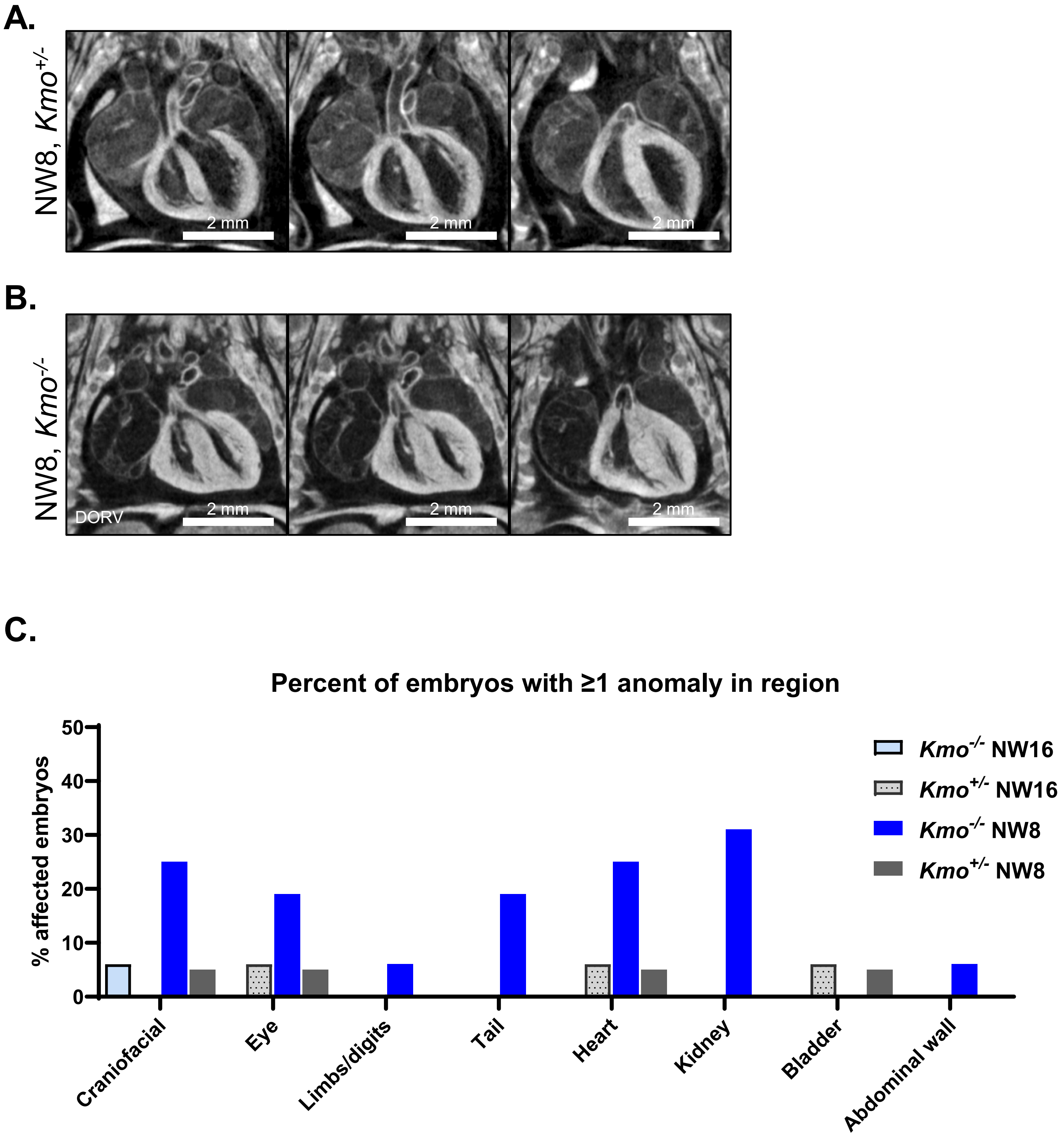


### Supplemental Figure 3

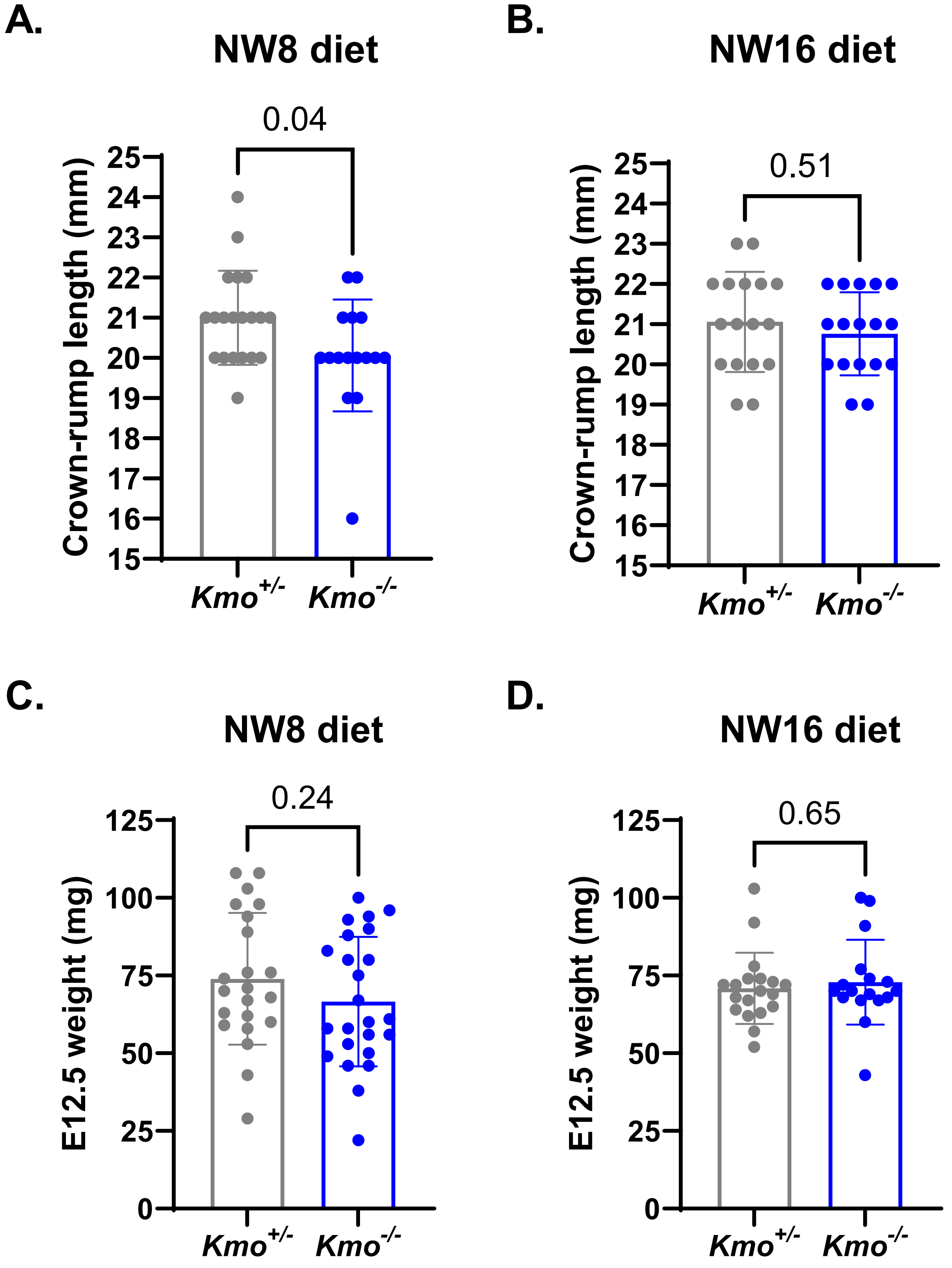


### Supplemental Figure 4

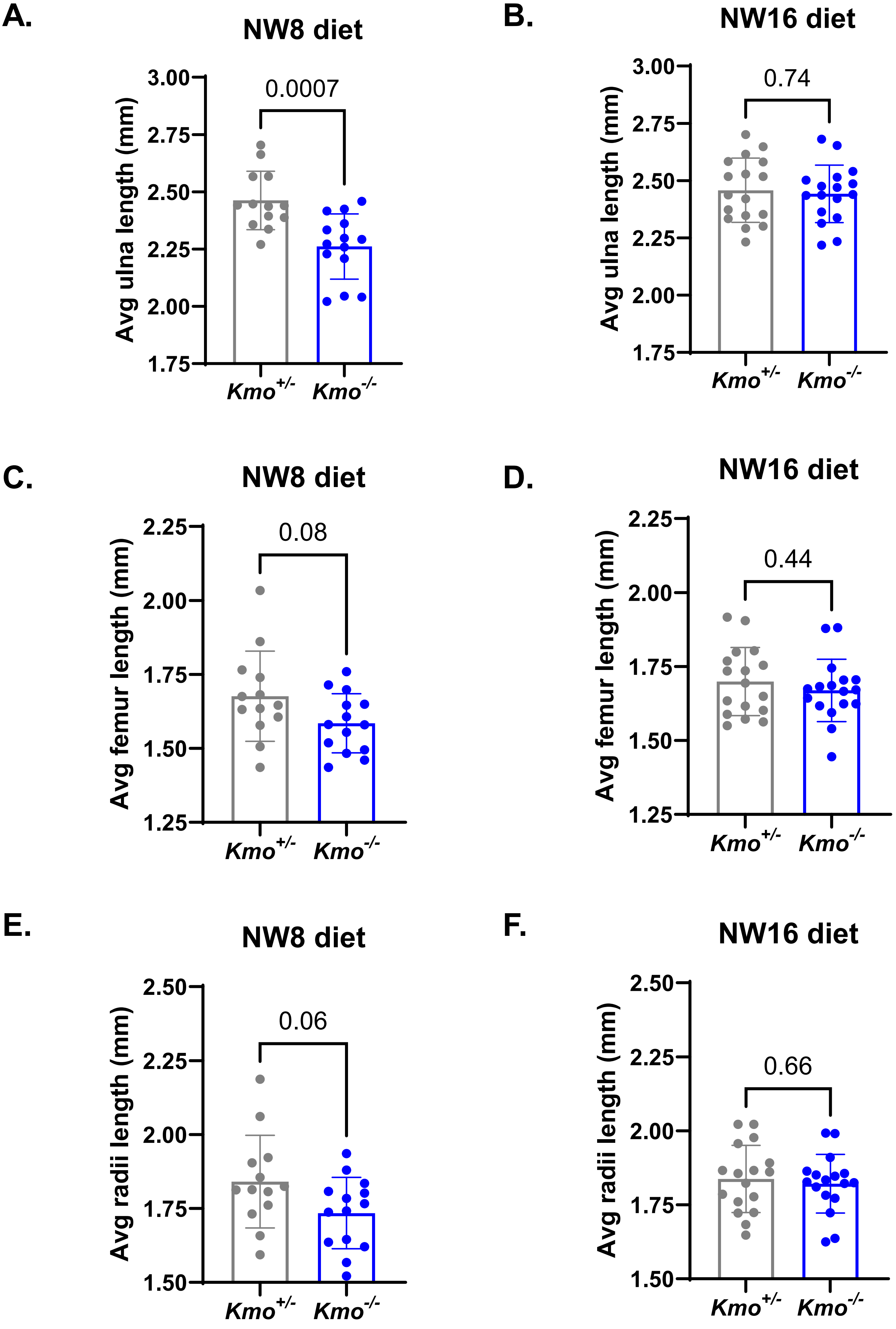


Supplemental Figure 5

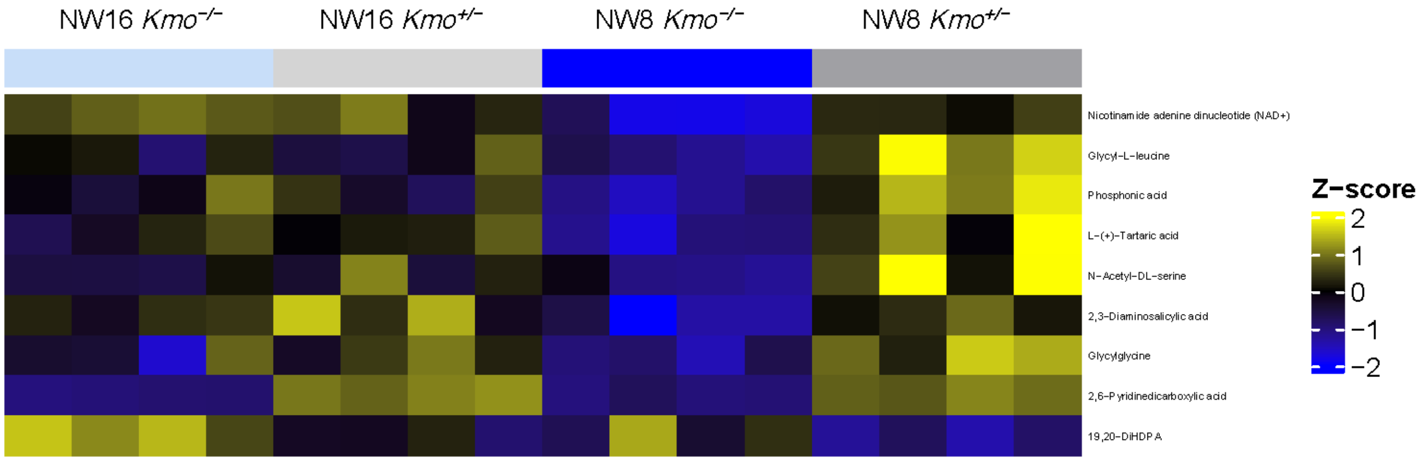


Metabolites significant for genotype effect

### Supplemental Figure 6

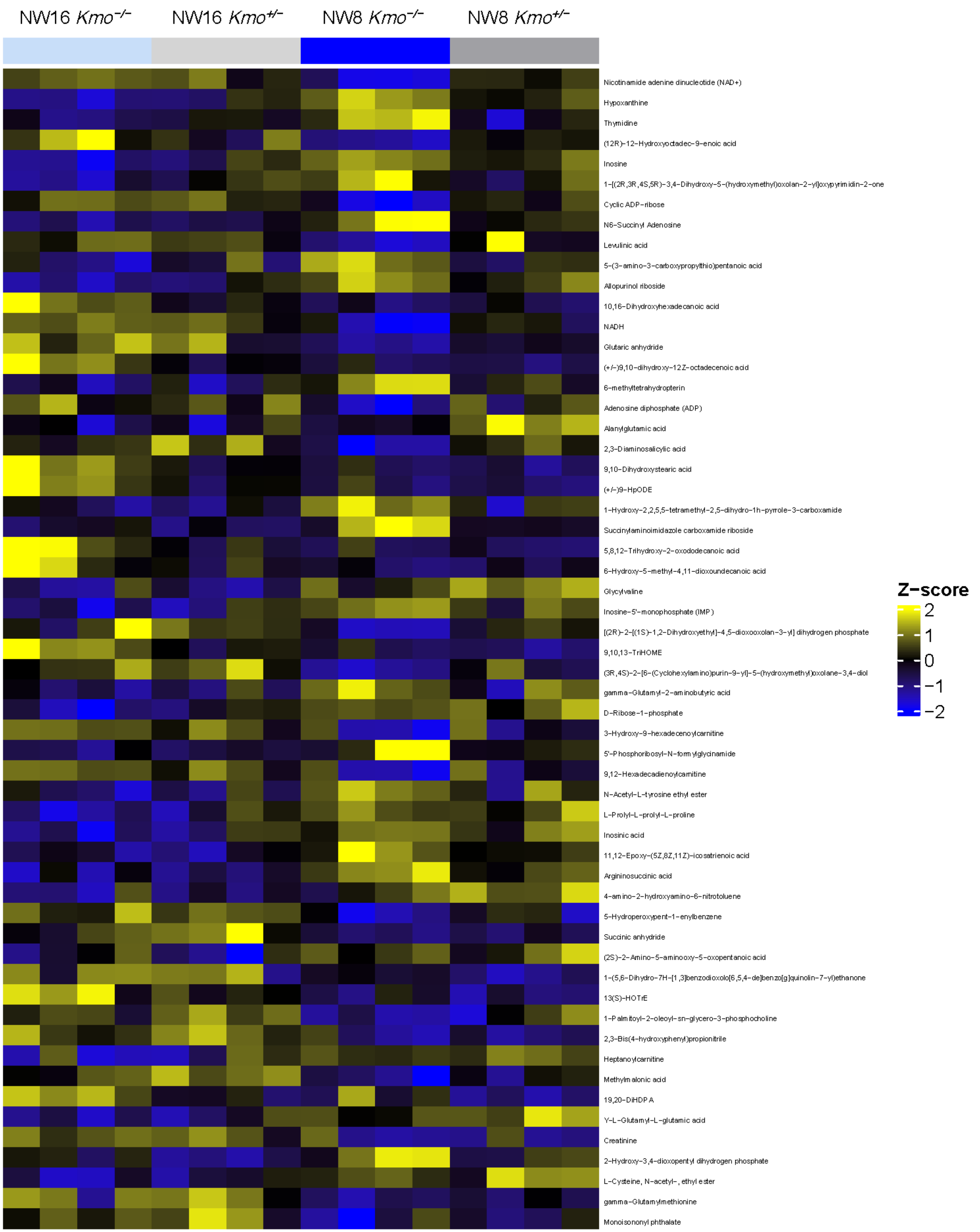


Metabolites significant for diet effect

### Supplemental Figure 7

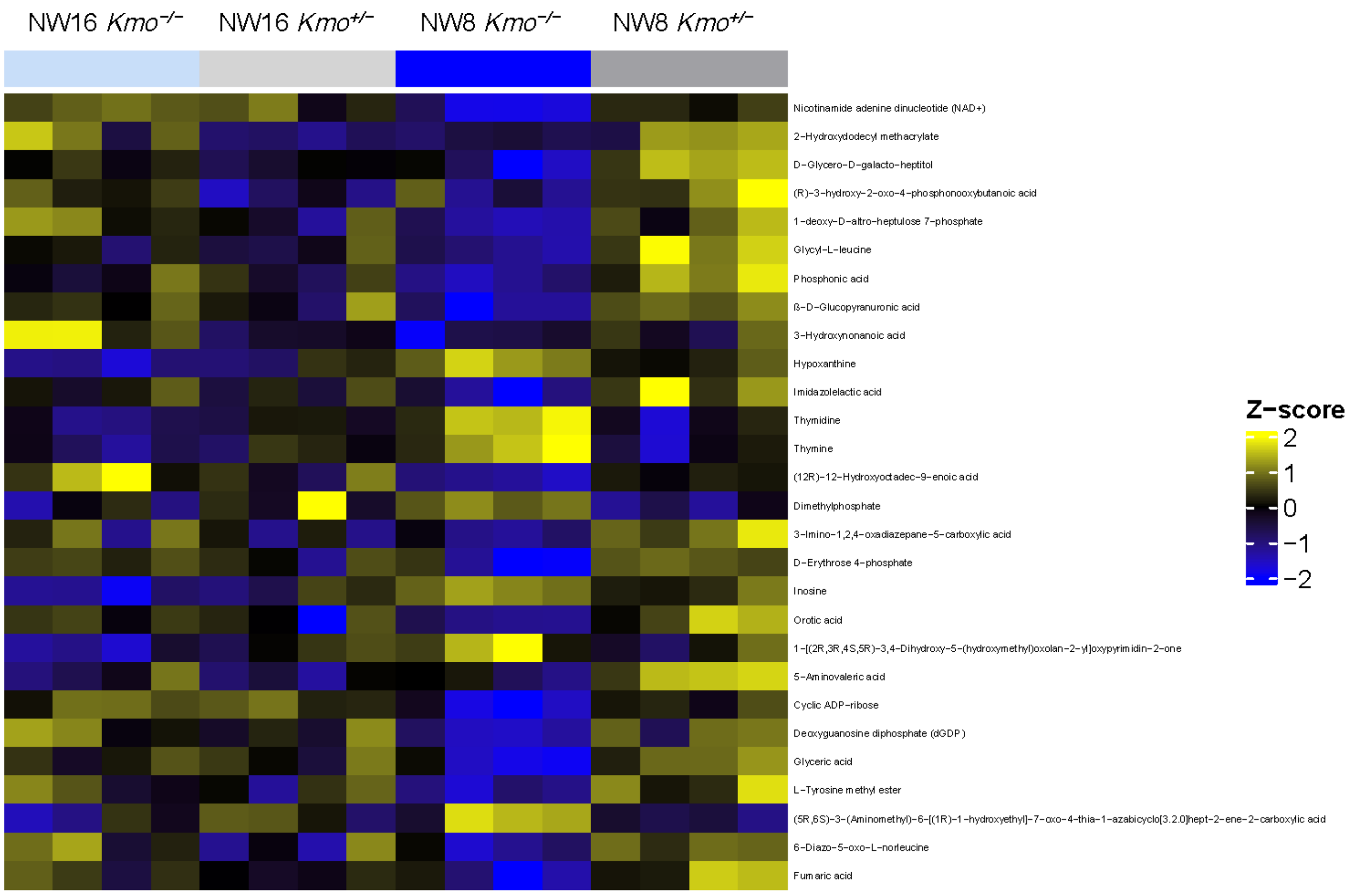


Metabolites significant for interaction (genotype × diet) effect

### Supplemental Figure 8

### 2-way ANOVA – significant interaction (FDR ≤ 0.25)

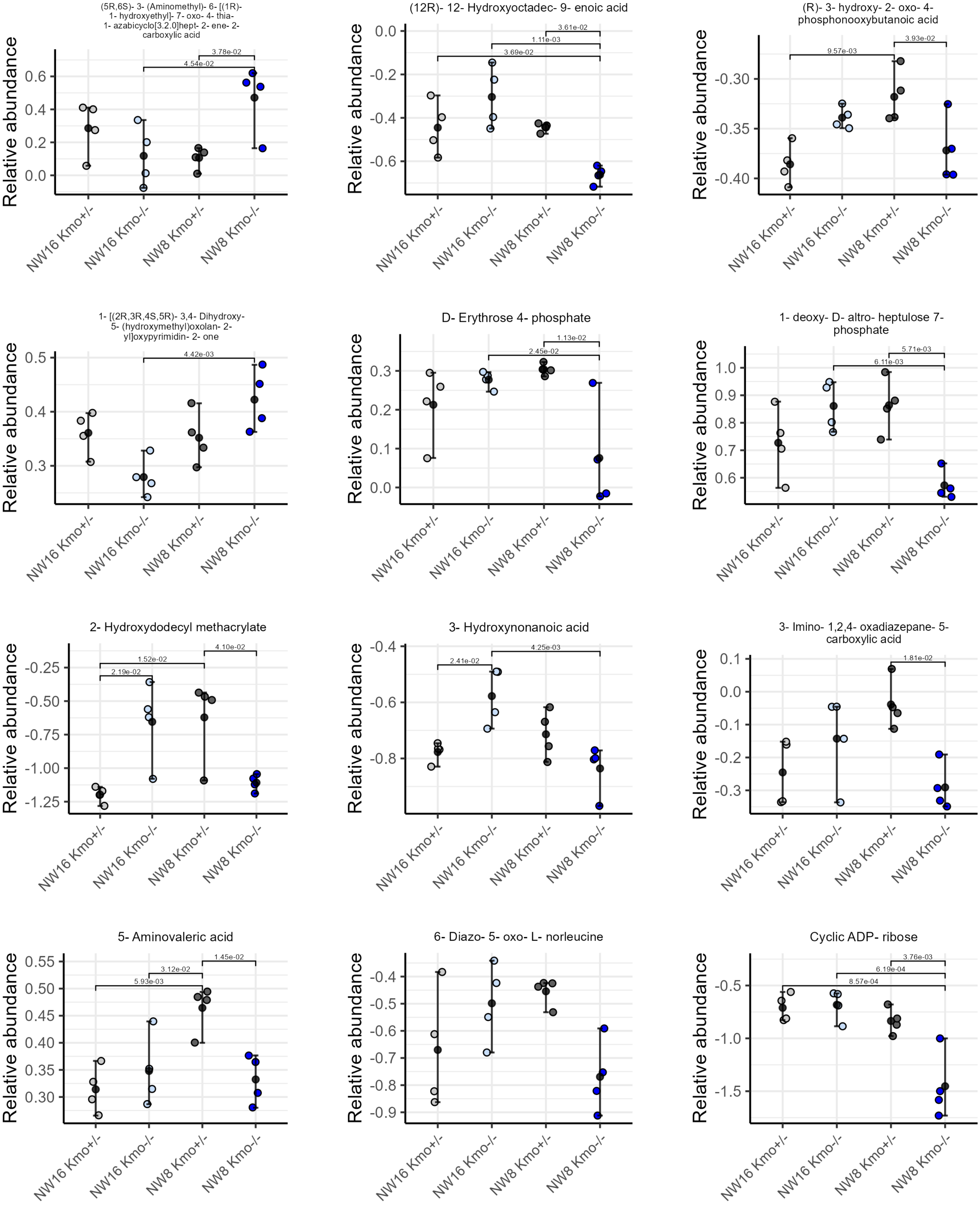


### Supplemental Figure 9

### 2-way ANOVA – significant interaction (FDR ≤ 0.25)

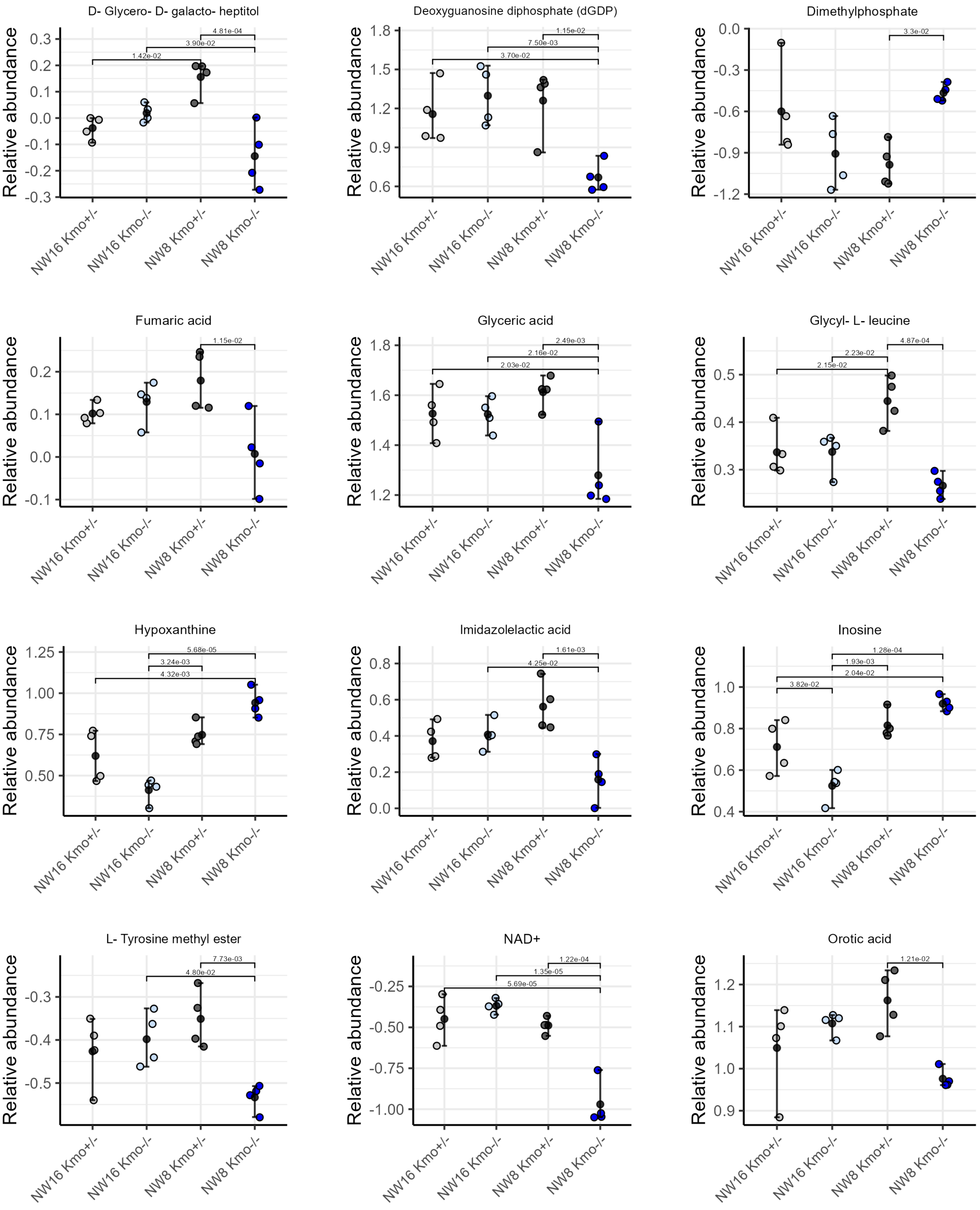


### Supplemental Figure 9, extended

### 2-way ANOVA – significant interaction (FDR ≤ 0.25)

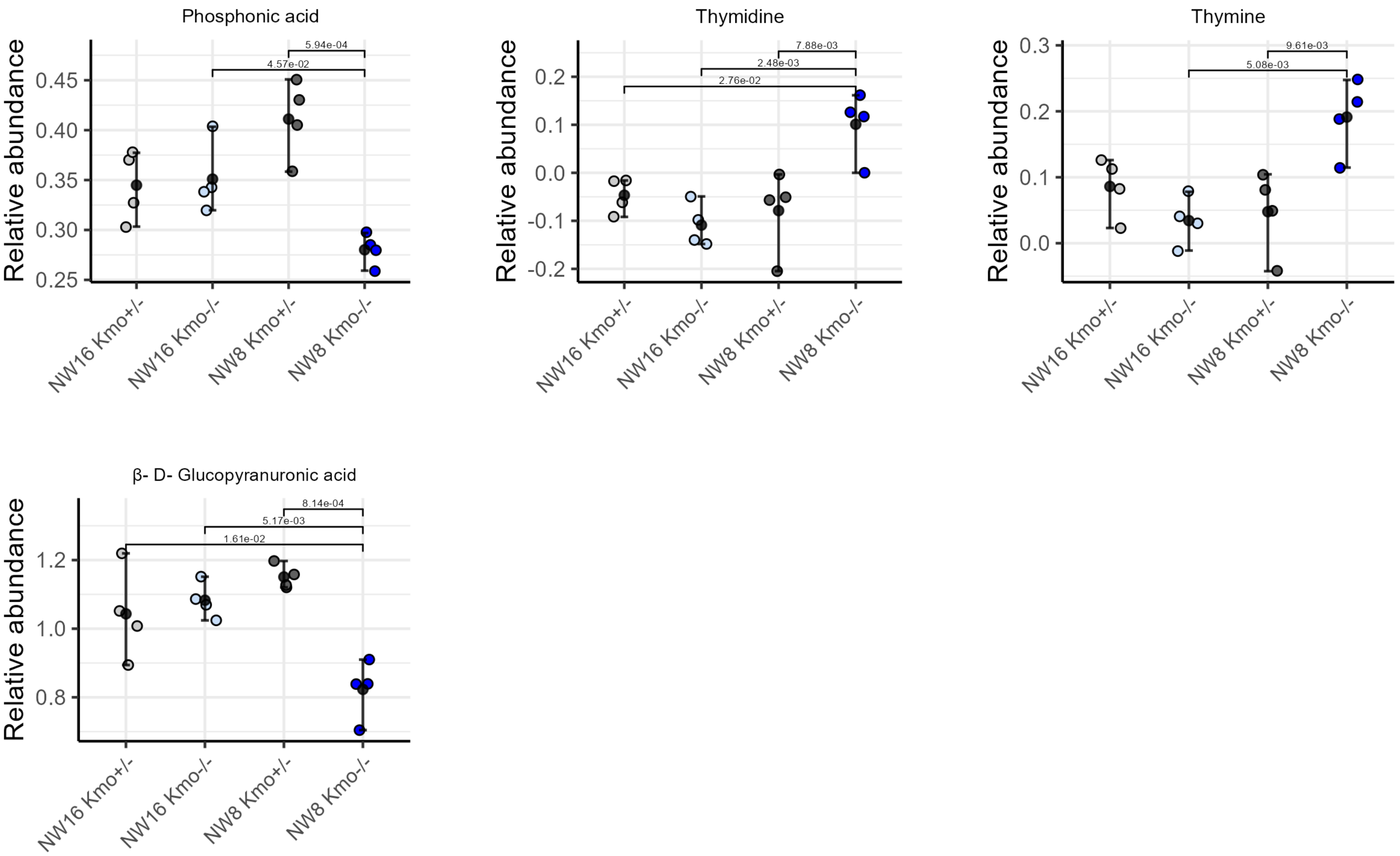


### Supplemental Figure 9, extended

**Supplemental Table 1**. Levels of metabolites of the NAD metabolome are altered in whole blood from the proband (II-2).

| **Metabolite** | **Individual and relation to proband** | | | | | | **Mean in unaffected individuals** | **Level in II-2 versus mean** |
|---|---|---|---|---|---|---|---|---|
| | I-1, Father | I-2, Mother | II-2, Proband | II-3, Sibling | Control female | Control male | | |
| **Kynurenine pathway, upstream of *KMO*** | | | | | | | | |
| TRP (μM) | 23.0 | 21.3 | 48.7 | 24.1 | 31.0 | 36.6 | 27.2 | 1.8 |
| KYN (μM) | 1.5 | 0.7 | 12.5 | 2.1 | 1.0 | 2.1 | 1.5 | 8.5 |
| KA (nM) | 65.8 | 30.9 | 597.8 | 28.8 | 21.5 | 40.5 | 37.5 | 15.9 |
| AA (nM) | 79.9 | 81.9 | 115.0 | 113.8 | 22.4 | 32.8 | 66.2 | 1.7 |
| **Kynurenine pathway, downstream of *KMO*** | | | | | | | | |
| XA (nM) | 5.3 | 4.5 | 2.1 | 1.0 | 3.8 | 13.6 | 5.6 | 0.4 |
| 3HAA (nM) | 22.1 | 17.8 | 19.6 | 15.3 | 15.7 | 26.6 | 19.5 | 1.0 |
| QA (nM) | 125.6 | 127.6 | 85.0 | 195.3 | 153.6 | 234.7 | 167.3 | 0.5 |

**Supplemental Table 2.** Adult *Kmo*$^{+/-}$ and *Kmo*$^{-/-}$ mice were observed at the expected mendelian ratio. E18.5 and E12.5 *Kmo*$^{+/-}$ and *Kmo*$^{-/-}$ embryos on the NW8 and NW16 diets were observed at the expected mendelian ratio. The sex distributions for E18.5 embryos on the NW8 and NW16 diets were also observed at the expected ratio.

**Adult/Liveborn mice**

| Genotype | Observed | Expected | $\chi^2$ p-value |
|---|---|---|---|
| ***Kmo***$^{-/-}$ | 162 | 155 | 0.610 |
| ***Kmo***$^{+/-}$ | 313 | 310 | |
| ***Kmo***$^{+/+}$ | 145 | 155 | |

**NW8 diet - E18.5 Embryos**

| Genotype | Observed | Expected | $\chi^2$ p-value |
|---|---|---|---|
| ***Kmo***$^{-/-}$ | 16 | 17.5 | 0.612 |
| ***Kmo***$^{+/-}$ | 19 | 17.5 | |
| **Total** | 35 | 35 | |
| **Sex** | **Observed** | **Expected** | **$\chi^2$ p-value** |
| **Male** | 20 | 17.5 | 0.398 |
| **Female** | 15 | 17.5 | |
| **Total** | 35 | 35 | |

**NW16 diet - E18.5 Embryos**

| Genotype | Observed | Expected | $\chi^2$ p-value |
|---|---|---|---|
| ***Kmo***$^{-/-}$ | 17 | 17 | 1 |
| ***Kmo***$^{+/-}$ | 17 | 17 | |
| **Total** | 34 | 34 | |
| **Sex** | **Observed** | **Expected** | **$\chi^2$ p-value** |
| **Male** | 19 | 17 | 0.493 |
| **Female** | 15 | 17 | |
| **Total** | 34 | 34 | |

**NW8 diet - E12.5 Embryos**

| Genotype | Observed | Expected | $\chi^2$ p-value |
|---|---|---|---|
| ***Kmo***$^{-/-}$ | 25 | 23.5 | 0.662 |
| ***Kmo***$^{+/-}$ | 22 | 23.5 | |
| **Total** | 47 | 47 | |

**NW16 diet - E12.5 Embryos**

| Genotype | Observed | Expected | $\chi^2$ p-value |
|---|---|---|---|
| ***Kmo***$^{-/-}$ | 17 | 18.5 | 0.662 |
| ***Kmo***$^{+/-}$ | 20 | 18.5 | |
| **Total** | 37 | 37 | |

****Tables for E12.5 embryos contain all collected embryos, not just embryos used for metabolomics or NAD measurements.

**Supplemental Table 3.** Genotyping, *Sry*, and RT-qPCR primers.

| Primer name | Sequence (5'-3') |
|---|---|
| Sry F | GAGAGCATGGAGGGCCAT |
| Sry R | CCACTCCTCTGTGACACT |
| Kmo E1 F | TGAATTGCGACACTCAGGGT |
| Kmo E1 R | GCATATTTCCATCAGAGCTCCC |
| Kmo K1 F | ACTGGTGTGGAGGTGAATACA |
| Kmo K1 R | ACAGCCGGTTAGCTTTCTGT |
| Kmo E2-4 F | GGAAGATATTCGCGTGGCTA |
| Kmo E2-4 R | CGATCTGATCTTCCAGACCAA |
| Kmo E9-10 F | GAAAGACTTCCAACGCGCAG |
| Kmo E9-10 R | CTGGGCAGGCAACAGAAAGA |
| Bact-F | TGTTACCAACTGGGACGACA |
| Bact-R | ACCTGGGTCATCTTTTCACG |

**Supplemental table 4.** Anatomical areas in murine embryos assessed for congenital anomalies by µCT.

| General region | Specific tissue/area |
|---|---|
| **Eye** | Eye |
| | Optic nerve |
| | Retina |
| **Craniofacial** | Brain (scalp swelling included) |
| | Nasal septum |
| | Nasal cavity |
| | Oral cavity |
| | Palate |
| | Tongue |
| | Epiglottis |
| **Throat** | Trachea |
| | Thyroid gland |
| | Submandibular gland |
| **Thoracic cavity** | Thymus |
| | Esophagus |
| | Heart |
| | Lung |
| | Pericardium cavity |
| | Brown fat |
| | Diaphragm |
| **Abdominopelvic cavity** | Peritoneal cavity |
| | Liver |
| | Stomach |
| | Pancreas |
| | Adrenal gland |
| | Kidney |
| | Intestine |
| | Spleen |
| | Bladder |
| | Ovary |
| | Testes |
| | Penis |
| | Rectum |

**Members of the Baylor College of Medicine Center for Precision Medicine Models (CPMM)**

| Names | Email address | Affiliation(s) |
|---|---|---|
| **Jason D Heaney** | | Department of Molecular and Human Genetics, Baylor College of Medicine, Houston Texas 77030; Texas Children's Hospital, Houston Texas 77030 |
| **Linday C Burrage** | | Department of Molecular and Human Genetics, Baylor College of Medicine, Houston Texas 77030 |
| **Denise G Lanza** | | Department of Molecular and Human Genetics, Baylor College of Medicine, Houston Texas 77030 |
| **Aleksander Milosavljevic** | | Department of Molecular and Human Genetics, Baylor College of Medicine, Houston Texas 77030 |
| **Matthew E Roth** | | Department of Molecular and Human Genetics, Baylor College of Medicine, Houston Texas 77030 |
| **Uma Ramamurthy** | | Office of Research Information Technology, Baylor College of Medicine, Houston Texas 77030 |
| **Jill A Rosenfeld** | | Department of Molecular and Human Genetics, Baylor College of Medicine, Houston Texas 77030 |
| **Shinya Yamamoto** | | Department of Molecular and Human Genetics, Baylor College of Medicine, Houston Texas 77030; Department of Neuroscience, Baylor College of Medicine, Houston Texas 77030; Jan and Dan Duncan Neurological Research Institute, Texas Children's Hospital, Houston Texas 77030 |
| **Michael F Wangler** | | Department of Molecular and Human Genetics, Baylor College of Medicine, Houston Texas 77030; Jan and Dan Duncan Neurological Research Institute, Texas Children's Hospital, Houston Texas 77030 |
| **Hugo J Bellen** | | Department of Molecular and Human Genetics, Baylor College of Medicine, Houston Texas 77030; Department of Neuroscience, BCM, Jan and Dan Duncan Neurological Research Institute, Texas Children's Hospital, Houston Texas 77030 |
| **Sandesh CS Nagamani** | | Department of Molecular and Human Genetics, Baylor College of Medicine, Houston Texas 77030 |
| **Seema R Lalani** | | Department of Molecular and Human Genetics, Baylor College of Medicine, Houston Texas 77030 |
| **Pengfei Liu** | | Department of Molecular and Human Genetics, Baylor College of Medicine, Houston Texas 77030 |
| **Ramin Darshoori** | | Department of Molecular and Human Genetics, Baylor College of Medicine, Houston Texas 77030 |
| **Chih-Wei Hsu** | | Department of Integrative Physiology, Baylor College of Medicine, Houston Texas 77030 |
| **Oguz Kanca** | | Department of Molecular and Human Genetics, Baylor College of Medicine, Houston Texas 77030; Jan and Dan Duncan Neurological Research Institute, Texas Children's Hospital, Houston Texas 77030 |

#### Members of the Undiagnosed Diseases Network (Version: 07.21.2026)

| Full Name | Affiliation | Component | Email |
|---|---|---|---|
| Alyssa A. Tran | Baylor College of Medicine | Clinical Site | |
| Arjun Tarakad | Baylor College of Medicine | Clinical Site | |
| Brendan H. Lee | Baylor College of Medicine | Clinical Site | |
| Carlos A. Bacino | Baylor College of Medicine | Clinical Site | |
| Christine M. Eng | Baylor College of Medicine | Sequencing Core | |
| Daryl A. Scott | Baylor College of Medicine | Clinical Site | |
| Elaine Seto | Baylor College of Medicine | Clinical Site | |
| Elizabeth Fieg | Baylor College of Medicine | Sequencing Core | |
| Fernando Scaglia | Baylor College of Medicine | Clinical Site | |
| Hongzheng Dai | Baylor College of Medicine | Clinical Site | |
| Hsiao-Tuan Chao | Baylor College of Medicine | Clinical Site | |
| Hugo J. Bellen | Baylor College of Medicine | Model Organisms Screening Center | |
| Ivan Chinn | Baylor College of Medicine | Clinical Site | |
| Jacqueline V. Chui | Baylor College of Medicine | Sequencing Core | |
| James P. Orengo | Baylor College of Medicine | Clinical Site | |
| Jared Sninsky | Baylor College of Medicine | Clinical Site | |
| Jill A. Rosenfeld | Baylor College of Medicine | Clinical Site | |
| Kim Worley | Baylor College of Medicine | Clinical Site | |
| Lauren Blieden | Baylor College of Medicine | Clinical Site | |
| Lindsay C. Burrage | Baylor College of Medicine | Clinical Site | |
| Michael F. Wangler | Baylor College of Medicine | Model Organisms Screening Center | |
| Monika Weisz Hubshman | Baylor College of Medicine | Clinical Site | |
| Nichole M. Owen | Baylor College of Medicine | Sequencing Core | |
| Pengfei Liu | Baylor College of Medicine | Sequencing Core | |
| Richard A. Lewis | Baylor College of Medicine | Clinical Site | |

| | | | |
|---|---|---|---|
| Ronit Marom | Baylor College of Medicine | Clinical Site | |
| Sandesh Nagamani | Baylor College of Medicine | Clinical Site | |
| Seema R. Lalani | Baylor College of Medicine | Clinical Site | |
| Shamika Ketkar | Baylor College of Medicine | Clinical Site | |
| Shinya Yamamoto | Baylor College of Medicine | Model Organisms Screening Center | |
| Tiphanie P. Vogel | Baylor College of Medicine | Clinical Site | |
| William J. Craigen | Baylor College of Medicine | Clinical Site | |
| Alan H. Beggs | Boston Children's Hospital | Clinical Site | |
| Ganesh Mochida | Boston Children's Hospital | Clinical Site | |
| Gerard T. Berry | Boston Children's Hospital | Clinical Site | |
| Ingrid A. Holm | Boston Children's Hospital | Clinical Site | |
| Lance H. Rodan | Boston Children's Hospital | Clinical Site | |
| Tina Truong | Boston Children's Hospital | Clinical Site | |
| Wendy Chung | Boston Children's Hospital | Clinical Site | |
| David Chiang | Brigham and Women's Hospital | Clinical Site | |
| Deepak A. Rao | Brigham and Women's Hospital | Clinical Site | |
| J. Carl Pallais | Brigham and Women's Hospital | Clinical Site | |
| Joseph Loscalzo | Brigham and Women's Hospital | Clinical Site | |
| Jose Abdenur | Children's Hospital of Orange County | Clinical Site | |
| Maija-Rikka Steenari | Children's Hospital of Orange County | Clinical Site | |
| Rebekah Barrick | Children's Hospital of Orange County | Clinical Site | |
| Richard Chang | Children's Hospital of Orange County | Clinical Site | |
| Cara Skraban | Children's Hospital of Philadelphia | Clinical Site | |
| Kathleen Sullivan | Children's Hospital of Philadelphia | Clinical Site | |
| Ramakrishnan Rajagopalan | Children's Hospital of Philadelphia | Clinical Site | |
| Rebecca Ganetzky | Children's Hospital of Philadelphia | Clinical Site | |
| Anne Slavotinek | Cincinnati Children's Hospital Medical Center | Clinical Site | |
| Christopher Mayhew | Cincinnati Children's Hospital Medical Center | Clinical Site | |
| Eneida Mendonca | Cincinnati Children's Hospital Medical Center | Clinical Site | |

| | | | |
|---|---|---|---|
| Ziyuan Guo | Cincinnati Children's Hospital Medical Center | Clinical Site | |
| Kelly Schoch | Duke University | Clinical Site | |
| Mohamad Mikati | Duke University | Clinical Site | |
| Nicole M. Walley | Duke University | Clinical Site | |
| Rebecca C. Spillmann | Duke University | Clinical Site | |
| Vandana Shashi | Duke University | Clinical Site | |
| Arjun K. Manrai | Harvard Medical School | Data Management Coordinating Center | |
| Cecilia Esteves | Harvard Medical School | Data Management Coordinating Center | |
| Emily Glanton | Harvard Medical School | Data Management Coordinating Center | |
| Isaac S. Kohane | Harvard Medical School | Data Management Coordinating Center | |
| Julie M. Johnson | Harvard Medical School | Data Management Coordinating Center | |
| Kimberly LeBlanc | Harvard Medical School | Data Management Coordinating Center | |
| Shilpa N. Kobren | Harvard Medical School | Data Management Coordinating Center | |
| Thomas A. Buckley | Harvard Medical School | Data Management Coordinating Center | |
| Ayuko Iverson | Icahn School of Medicine at Mount Sinai | Clinical Site | |
| Bruce Gelb | Icahn School of Medicine at Mount Sinai | Clinical Site | |
| Charlotte Cunningham-Rundles | Icahn School of Medicine at Mount Sinai | Clinical Site | |
| Eric Gayle | Icahn School of Medicine at Mount Sinai | Clinical Site | |
| Joanna Jen | Icahn School of Medicine at Mount Sinai | Clinical Site | |
| Louise Bier | Icahn School of Medicine at Mount Sinai | Clinical Site | |

| | | | |
|---|---|---|---|
| Mafalda Barbosa | Icahn School of Medicine at Mount Sinai | Clinical Site | |
| Manisha Balwani | Icahn School of Medicine at Mount Sinai | Clinical Site | |
| Mariya Shadrina | Icahn School of Medicine at Mount Sinai | Clinical Site | |
| Rachel Evard | Icahn School of Medicine at Mount Sinai | Clinical Site | |
| Rory M.C. Abrams | Icahn School of Medicine at Mount Sinai | Clinical Site | |
| Saskia Shuman | Icahn School of Medicine at Mount Sinai | Clinical Site | |
| Brett H. Graham | Indiana University School of Medicine | Clinical Site | |
| Erin Conboy | Indiana University School of Medicine | Clinical Site | |
| Francesco Vetrini | Indiana University School of Medicine | Clinical Site | |
| Kayla M. Treat | Indiana University School of Medicine | Clinical Site | |
| Khurram Liaqat | Indiana University School of Medicine | Clinical Site | |
| Lili Mantcheva | Indiana University School of Medicine | Clinical Site | |
| Stephanie M. Ware | Indiana University School of Medicine | Clinical Site | |
| Elizabeth Wohler | Johns Hopkins University | Clinical Site | |
| Julie Hoover-Fong | Johns Hopkins University | Clinical Site | |
| Kathleen Page | Johns Hopkins University | Clinical Site | |
| Matthew Robinson | Johns Hopkins University | Clinical Site | |
| Nara Sobreira | Johns Hopkins University | Clinical Site | |
| P Dane Witmer | Johns Hopkins University | Clinical Site | |
| Paul Auwaerter | Johns Hopkins University | Clinical Site | |
| Winston Timp | Johns Hopkins University | Clinical Site | |
| Yuka Manabe | Johns Hopkins University | Clinical Site | |
| David A. Sweetser | Massachusetts General Hospital | Clinical Site | |
| Frances High | Massachusetts General Hospital | Clinical Site | |
| Lauren C. Briere | Massachusetts General Hospital | Clinical Site | |
| Melissa Walker | Massachusetts General Hospital | Clinical Site | |
| Breanna Mitchell | Mayo Clinic | Clinical Site | |
| Brendan C. Lanpher | Mayo Clinic | Clinical Site | |
| Devin Oglesbee | Mayo Clinic | Clinical Site | |

| | | | |
|---|---|---|---|
| Eric Klee | Mayo Clinic | Clinical Site | |
| Erin A. Wishart | Mayo Clinic | Clinical Site | |
| Filippo Pinto e Vairo | Mayo Clinic | Clinical Site | |
| Kahlen Darr | Mayo Clinic | Clinical Site | |
| Laura C. Duncan | Mayo Clinic | Clinical Site | |
| Lindsay Mulvihill | Mayo Clinic | Clinical Site | |
| Lisa Schimmenti | Mayo Clinic | Clinical Site | |
| Queenie Tan | Mayo Clinic | Clinical Site | |
| Abdul Elkadri | Medical College of Wisconsin and Children's Wisconsin | Clinical Site | |
| Brett Bordini | Medical College of Wisconsin and Children's Wisconsin | Clinical Site | |
| Donald Basel | Medical College of Wisconsin and Children's Wisconsin | Clinical Site | |
| James Verbsky | Medical College of Wisconsin and Children's Wisconsin | Clinical Site | |
| Julie McCarrier | Medical College of Wisconsin and Children's Wisconsin | Clinical Site | |
| Michael Muriello | Medical College of Wisconsin and Children's Wisconsin | Clinical Site | |
| Michael T. Zimmermann | Medical College of Wisconsin and Children's Wisconsin | Clinical Site | |
| Chantale Branson | Morehouse School of Medicine | Data Management Coordinating Center | |
| Herman Taylor | Morehouse School of Medicine | Data Management Coordinating Center | |
| Kisha J Young MD PhD | Morehouse School of Medicine | Clinical Site | |
| Rakale C. Quarells | Morehouse School of Medicine | Data Management Coordinating Center | |
| Andrea Gropman | National Institutes of Health | Clinical Site | |
| Barbara N. Pusey Swerdzewski | National Institutes of Health | Clinical Site | |
| Ben Afzali | National Institutes of Health | Clinical Site | |
| Ben Solomon | National Institutes of Health | Clinical Site | |
| Camilo Toro | National Institutes of Health | Clinical Site | |

| | | | |
|---|---|---|---|
| Colleen E. Wahl | National Institutes of Health | Clinical Site | |
| Cynthia J. Tifft | National Institutes of Health | Clinical Site | |
| David R. Adams | National Institutes of Health | Clinical Site | |
| Donna Novacic | National Institutes of Health | Clinical Site | |
| Elizabeth A. Burke | National Institutes of Health | Clinical Site | |
| Ellen F. Macnamara | National Institutes of Health | Clinical Site | |
| Heidi Wood | National Institutes of Health | Clinical Site | |
| Jiayu Fu | National Institutes of Health | Clinical Site | |
| Joie Davis | National Institutes of Health | Clinical Site | |
| Leoyklang Petcharet | National Institutes of Health | Clinical Site | |
| Lynne A. Wolfe | National Institutes of Health | Clinical Site | |
| Margaret Delgado | National Institutes of Health | Clinical Site | |
| Maria T. Acosta | National Institutes of Health | Clinical Site | |
| Marie Morimoto | National Institutes of Health | Clinical Site | |
| Marla Sabaii | National Institutes of Health | Clinical Site | |
| May Christine V. Malicdan | National Institutes of Health | Clinical Site | |
| Neil Hanchard | National Institutes of Health | Clinical Site | |
| Orpa Jean-Marie | National Institutes of Health | Clinical Site | |
| Precilla D'Souza | National Institutes of Health | Clinical Site | precilla.d' |
| Valerie V. Maduro | National Institutes of Health | Clinical Site | |
| Wendy Introne | National Institutes of Health | Clinical Site | |
| William A. Gahl | National Institutes of Health | Clinical Site | |
| Yan Huang | National Institutes of Health | Clinical Site | |
| Vaidehi Jobanputra | New York Genome Center | Clinical Site | |
| Chun-Hung Chan | Sanford Research | Clinical Site | |
| D Isum Ward | Sanford Research | Clinical Site | |
| Debbie Figueroa | Sanford Research | Clinical Site | |
| Francisco Bustos | Sanford Research | Clinical Site | |
| Jason Schend | Sanford Research | Clinical Site | |
| Jennifer Morgan | Sanford Research | Clinical Site | |
| Megan Bell | Sanford Research | Clinical Site | |
| Miranda Leitheiser | Sanford Research | Clinical Site | |
| Mohamad Saifeddine | Sanford Research | Clinical Site | |

| | | | |
|---|---|---|---|
| Paul Berger | Sanford Research | Clinical Site | |
| Rachel Li | Sanford Research | Clinical Site | |
| Taylor Beagle | Sanford Research | Clinical Site | |
| Emily Shelkowitz | Seattle Children's Hospital | Clinical Site | |
| Eric Allenspach | Seattle Children's Hospital | Clinical Site | |
| Katrina Dipple | Seattle Children's Hospital | Clinical Site | |
| Seth Perlman | Seattle Children's Hospital | Clinical Site | |
| Beth A. Martin | Stanford University | Clinical Site | |
| Chloe M. Reuter | Stanford University | Clinical Site | |
| Dena R. Matalon | Stanford University | Clinical Site | |
| Devon Bonner | Stanford University | Clinical Site | |
| Euan A. Ashley | Stanford University | Data Management Coordinating Center | |
| Hector Rodrigo Mendez | Stanford University | Clinical Site | |
| Holly K. Tabor | Stanford University | Clinical Site | |
| Jacinda B. Sampson | Stanford University | Clinical Site | |
| Jason Hom | Stanford University | Clinical Site | |
| Jennefer N. Kohler | Stanford University | Clinical Site | |
| Jennifer Schymick | Stanford University | Clinical Site | |
| Jonathan A. Bernstein | Stanford University | Clinical Site | |
| Kevin S. Smith | Stanford University | Clinical Site | |
| Laura Keehan | Stanford University | Clinical Site | |
| Laurens Wiel | Stanford University | Clinical Site | |
| Matthew T. Wheeler | Stanford University | Clinical Site | |
| Meghan C. Halley | Stanford University | Clinical Site | |
| Mia Levanto | Stanford University | Clinical Site | |
| Paul G. Fisher | Stanford University | Clinical Site | |
| Rachel A. Ungar | Stanford University | Clinical Site | |
| Raquel L. Alvarez | Stanford University | Clinical Site | |
| Shruti Marwaha | Stanford University | Clinical Site | |
| Sky Kim | Stanford University | Clinical Site | |
| Sophia Adelson | Stanford University | Clinical Site | |
| Stephen B Montgomery | Stanford University | Clinical Site | |
| Suha Bachir | Stanford University | Clinical Site | |

| | | | |
|---|---|---|---|
| Tanner D Jensen | Stanford University | Clinical Site | |
| Taylor Maurer | Stanford University | Clinical Site | |
| Terra R. Coakley | Stanford University | Clinical Site | |
| Danielle Carnival | Undiagnosed Diseases Network Foundation | Undiagnosed Diseases Network Foundation | |
| Sarah Marshall | Undiagnosed Diseases Network Foundation | Undiagnosed Diseases Network Foundation | |
| Aleksandra Foksinska | University of Alabama at Birmingham | Data Management Coordinating Center | |
| Andrew B. Crouse | University of Alabama at Birmingham | Data Management Coordinating Center | |
| Anna Hurst | University of Alabama at Birmingham | Clinical Site | |
| Brandon M Wilk | University of Alabama at Birmingham | Clinical Site | |
| Bruce R Korf | University of Alabama at Birmingham | Clinical Site | |
| C-H. Wilfred Wu | University of Alabama at Birmingham | Clinical Site | |
| Elizabeth A Worthey | University of Alabama at Birmingham | Clinical Site | |
| Kaitlin Callaway | University of Alabama at Birmingham | Clinical Site | |
| Martin Rodriguez | University of Alabama at Birmingham | Clinical Site | |
| Matthew Might | University of Alabama at Birmingham | Data Management Coordinating Center | |
| Pongtawat Lertwilaiwittaya | University of Alabama at Birmingham | Clinical Site | |
| Reaford Blackburn | University of Alabama at Birmingham | Clinical Site | |
| Teneasha Washington | University of Alabama at Birmingham | Clinical Site | |
| William E. Byrd | University of Alabama at Birmingham | Data Management Coordinating Center | |
| Albert R. La Spada | University of California, Irvine | Clinical Site | |
| Changrui Xiao | University of California, Irvine | Clinical Site | |
| Christina Nyugen | University of California, Irvine | Clinical Site | |
| Emmanuèle C. Délot | University of California, Irvine | Clinical Site | |
| Eric Vilain | University of California, Irvine | Clinical Site | |
| Fuki M. Hisama | University of California, Irvine | Clinical Site | |
| Giovanna S. Manzano | University of California, Irvine | Clinical Site | |
| Sanaz Attaripour | University of California, Irvine | Clinical Site | |
| Tahseen Mozaffar | University of California, Irvine | Clinical Site | |

| | | | |
|---|---|---|---|
| Yongen Chang | University of California, Irvine | Clinical Site | |
| Alden Huang | University of California, Los Angeles | Clinical Site | |
| Andres Vargas | University of California, Los Angeles | Clinical Site | |
| Brent L. Fogel | University of California, Los Angeles | Clinical Site | |
| Daniela Nasif | University of California, Los Angeles | Clinical Site | |
| George Carvalho | University of California, Los Angeles | Clinical Site | |
| Julian A. Martínez-Agosto | University of California, Los Angeles | Clinical Site | |
| Layal F. Abi Farraj | University of California, Los Angeles | Clinical Site | |
| Manish J. Butte | University of California, Los Angeles | Clinical Site | |
| Martin G. Martin | University of California, Los Angeles | Clinical Site | |
| Naghmeh Dorrani | University of California, Los Angeles | Clinical Site | |
| Rosario I. Corona | University of California, Los Angeles | Clinical Site | |
| Stanley F. Nelson | University of California, Los Angeles | Clinical Site | |
| Carson A. Smith | University of Miami | Clinical Site | |
| Deborah Barbouth | University of Miami | Clinical Site | |
| Guney Bademci | University of Miami | Clinical Site | |
| Joanna M. Gonzalez | University of Miami | Clinical Site | |
| Kumarie Latchman | University of Miami | Clinical Site | |
| LéShon Peart | University of Miami | Clinical Site | |
| Mustafa Tekin | University of Miami | Clinical Site | |
| Nicholas Borja | University of Miami | Clinical Site | |
| Stephan Zuchner | University of Miami | Clinical Site | |
| Stephanie Bivona | University of Miami | Clinical Site | |
| Willa Thorson | University of Miami | Clinical Site | |
| Monte Westerfield | University of Oregon | Model Organisms Screening Center | |
| Anna Raper | University of Pennsylvania | Clinical Site | |
| Daniel J. Rader | University of Pennsylvania | Clinical Site | |
| Giorgio Sirugo | University of Pennsylvania | Clinical Site | |
| Aaron Quinlan | University of Utah | Clinical Site | |
| Alistair Ward | University of Utah | Clinical Site | |
| Ashley Andrews | University of Utah | Clinical Site | |
| Corrine K. Welt | University of Utah | Clinical Site | |

| | | | |
|---|---|---|---|
| Dave Viskochil | University of Utah | Clinical Site | |
| Erin E. Baldwin | University of Utah | Clinical Site | |
| Gabor Marth | University of Utah | Data Management Coordinating Center | |
| John Carey | University of Utah | Clinical Site | |
| Lorenzo Botto | University of Utah | Clinical Site | |
| Matt Velinder | University of Utah | Clinical Site | |
| Nicola Longo | University of Utah | Clinical Site | |
| Paolo Moretti | University of Utah | Clinical Site | |
| Pinar Bayrak-Toydemir | University of Utah | Clinical Site | |
| Rebecca Overbury | University of Utah | Clinical Site | |
| Rong Mao | University of Utah | Clinical Site | |
| Russell Butterfield | University of Utah | Clinical Site | |
| Steven Boyden | University of Utah | Clinical Site | |
| Thomas J. Nicholas | University of Utah | Clinical Site | |
| Andrew Stergachis | University of Washington | Clinical Site | |
| Annelise Mah-Som | University of Washington | Clinical Site | |
| Danny E. Miller | University of Washington | Clinical Site | |
| Elisabeth Rosenthal | University of Washington | Clinical Site | |
| Elizabeth Blue | University of Washington | Clinical Site | |
| Elsa Balton | University of Washington | Clinical Site | |
| Gail P. Jarvik | University of Washington | Clinical Site | |
| Ghayda Mirzaa | University of Washington | Clinical Site | |
| Ian Glass | University of Washington | Clinical Site | |
| Kathleen A. Leppig | University of Washington | Clinical Site | |
| Mark Wener | University of Washington | Clinical Site | |
| Martha Horike-Pyne | University of Washington | Clinical Site | |
| Michael Bamshad | University of Washington | Clinical Site | |
| Peter Byers | University of Washington | Clinical Site | |
| Runjun Kumar | University of Washington | Clinical Site | |
| Sirisak Chanprasert | University of Washington | Clinical Site | |
| Virginia Sybert | University of Washington | Clinical Site | |
| Wendy Raskind | University of Washington | Clinical Site | |
| Bryn D. Webb | University of Wisconsin Madison | Clinical Site | |

| | | | |
|---|---|---|---|
| Kim M. Keppler-Noreuil | University of Wisconsin Madison | Clinical Site | |
| M. Stephen Meyn | University of Wisconsin Madison | Clinical Site | |
| Qiang Chang | University of Wisconsin Madison | Clinical Site | |
| Alyson Krokosky | Vanderbilt University Medical Center | Clinical Site | |
| Ashley McMinn | Vanderbilt University Medical Center | Clinical Site | |
| Austin Herbert | Vanderbilt University Medical Center | Clinical Site | |
| Cathy Shyr | Vanderbilt University Medical Center | Clinical Site | |
| Eric Gamazon | Vanderbilt University Medical Center | Clinical Site | |
| John A. Phillips III | Vanderbilt University Medical Center | Clinical Site | |
| Joy D. Cogan | Vanderbilt University Medical Center | Clinical Site | |
| Kimberly Ezell | Vanderbilt University Medical Center | Clinical Site | |
| Lakshitha Perera | Vanderbilt University Medical Center | Clinical Site | |
| Lisa Bastarache | Vanderbilt University Medical Center | Clinical Site | |
| Lynette Rives | Vanderbilt University Medical Center | Clinical Site | |
| Mary Koziura | Vanderbilt University Medical Center | Clinical Site | |
| Rizwan Hamid | Vanderbilt University Medical Center | Clinical Site | |
| Thomas Cassini | Vanderbilt University Medical Center | Clinical Site | |
| Alex Paul | Washington University in St. Louis | Clinical Site | |
| Dana Kiley | Washington University in St. Louis | Clinical Site | |
| Daniel Wegner | Washington University in St. Louis | Clinical Site | |
| Dustin Baldridge | Washington University in St. Louis | Model Organisms Screening Center | |
| F. Sessions Cole | Washington University in St. Louis | Data Management Coordinating Center | |
| Jennifer Wambach | Washington University in St. Louis | Clinical Site | |
| Jimann Shin | Washington University in St. Louis | Model Organisms Screening Center | |
| Jonathan Baker | Washington University in St. Louis | Clinical Site | |
| Kathleen A. Sisco | Washington University in St. Louis | Clinical Site | |
| Lilianna Solnica-Krezel | Washington University in St. Louis | Model Organisms Screening Center | |
| Patricia Dickson | Washington University in St. Louis | Clinical Site | |
| Robert McKinstry | Washington University in St. Louis | Clinical Site | |
| Stephen C. Pak | Washington University in St. Louis | Model Organisms Screening Center | |

| | | | |
|---|---|---|---|
| Timothy Schedl | Washington University in St. Louis | Model Organisms Screening Center | |
| Fan Ma | Yale University | Undiagnosed Diseases Network Foundation | |
| Haley Xiaohe Zhang | Yale University | Undiagnosed Diseases Network Foundation | |
| Julieta Bonvin Sallago | Yale University | Clinical Site | |
| Lauren Jeffries | Yale University | Clinical Site | |
| Majid Farhadloo | Yale University | Clinical Site | |
| María José Ortuño Romero | Yale University | Clinical Site | |
| Monkol Lek | Yale University | Clinical Site | |
| Teodoro Jerves Serrano | Yale University | Clinical Site | |
| Yong-Hui Jiang | Yale University | Clinical Site | |
